# The Incidence and Cumulative Risk of Major Surgery in Older Persons in the United States

**DOI:** 10.1101/2020.11.26.20239228

**Authors:** Robert D. Becher, Brent Vander Wyk, Linda Leo-Summers, Mayur M. Desai, Thomas M. Gill

**Author notes:** **For Correspondence:** Robert D. Becher, MD, MS, Division of General Surgery, Trauma, and Surgical Critical Care, Department of Surgery, Yale School of Medicine, 330 Cedar Street, BB-310, New Haven, CT 06520.

## Abstract

**Importance:** As the population of the United States (US) ages, there is considerable interest in ensuring safe and high-quality surgical care for older persons. Yet, valid, generalizable data on the occurrence of major surgery in the geriatric population are sparse.

**Objective:** To estimate the incidence and cumulative risk of major surgery in older persons over a 5-year period and evaluate how these estimates differ according to demographic and geriatric characteristics.

**Design:** Prospective longitudinal study.

**Setting:** Continental US from 2011 to 2016.

**Participants:** 5,571 community-living fee-for-service Medicare beneficiaries, aged 65+, from the National Health and Aging Trends Study (NHATS).

**Main Outcomes and Measures:** Major surgeries were identified through linkages with data from the Centers for Medicare & Medicaid Services. Data on frailty and dementia were obtained from the baseline NHATS assessment.

**Results:** The nationally-representative incidence of major surgery per 100 person-years was 8.8 (95% confidence interval [CI], 8.2-9.5), with estimates of 5.2 (95% CI, 4.7-5.7) and 3.7 (95% CI, 3.3-4.1) for elective and non-elective surgeries. The adjusted incidence of major surgery peaked at 10.8 (95% CI, 9.4-12.4) in persons 75-79 years, increased from 6.6 (95% CI, 5.8-7.5) in the non-frail group to 10.3 (95% CI, 8.9-11.9) in the frail group, and was similar by sex (males 8.6 [95% CI, 7.7-9.6]; females 8.3 [95% CI, 7.5-9.1]) and dementia (no 8.6 [95% CI, 7.9-9.3]; possible 7.8 [95% CI, 6.3-9.6]; probable 8.1 [95% CI, 6.7-9.9]). The 5-year cumulative risk of major surgery was 13.8% (95% CI, 12.2%-15.5%), representing nearly 5 million unique older persons (4,958,048 [95% CI, 4,345,342-5,570,755]), including 12.1% (95% CI, 9.5%-14.6%) in persons 85-89 years, 9.1% (95% CI, 7.2%-11.0%) in those ≥90 years, 12.1% (95% CI, 9.9%-14.4%) in those with frailty, and 12.4% (95% CI, 9.8%-15.0%) in those with probable dementia.

**Conclusions and Relevance:** Major surgery is a common event in the lives of community-living older persons, including high-risk vulnerable subgroups such as the oldest old, those with frailty or dementia, and those undergoing non-elective surgery. The burden of major surgery in older Americans will add to the challenges ahead for the US health care system in our aging society.

**KEY POINTS:** *Question:* What is the incidence and cumulative risk of major surgery in older persons in the United States?

*Findings:* In this prospective longitudinal study, data from 5,571 community-living fee-for-service Medicare beneficiaries were used to calculate nationally-representative estimates for the incidence and cumulative risk of major surgery over a 5-year period. Nearly 9 major surgeries were performed annually for every 100 older persons, and more than 1 in 7 Medicare beneficiaries underwent a major surgery over 5 years, representing nearly 5 million unique older persons.

*Meaning:* Major surgery is a common event in the lives of community-living older persons.

---

The projected growth of the geriatric population has been called the most significant demographic trend in the history of the United States (US).^1,2^ The number of persons aged 65 years or older is expected to double between now and 2060, from 46 to 98 million.^1,3^ While the expanding geriatric population will affect all areas of medicine, one field that will be especially impacted is surgery.^4^

As the US population ages, the number of older persons who will require surgical intervention will increase substantially.^5–7^ Because advancing age, frailty, dementia, and other geriatric-specific conditions are all risk factors for adverse outcomes after surgery,^8–16^ there is considerable interest in ensuring safe and high-quality surgical care for older persons.^17^ Despite this demographic imperative, valid and generalizable data on the epidemiology of major surgery in older Americans are lacking. Estimates of surgery in the geriatric population are outdated, extrapolated from state-level assessments, and based on administrative discharge data. In addition, prior studies have employed a broad definition of a surgical procedure and have not included values for clinically relevant subgroups, such as those who are frail or cognitively impaired.^4,5,18–23^ Reliable, up-to-date national estimates of the occurrence of major surgery in older persons are needed to inform public health policy and planning, enhance the accuracy of medical and surgical needs assessments by academic, health care, and commercial institutions, and identify opportunities to intervene in clinical practice.

The objectives of the current study were two-fold: first, to estimate the incidence and cumulative risk of major surgery in older persons in the US, across the spectrum of surgical disciplines, including both elective and non-elective operations; and second, to evaluate how these estimates differ according to key demographic and geriatric characteristics, including frailty and dementia. To accomplish these objectives, we used data from the National Health and Aging Trends Study^24^ (NHATS), linked to data from the Centers for Medicare & Medicaid Services (CMS).

## METHODS

### Data Sources

NHATS is a prospective, nationally-representative longitudinal study of Medicare beneficiaries.^24^ On September 30, 2010, NHATS drew a random sample of persons 65 years or older living in the contiguous US (excluding Alaska, Hawaii, and Puerto Rico) from the Medicare enrollment file, with oversampling of non-Hispanic blacks and those 90 years or older.^25^ Baseline (Round 1) interviews, completed from May through November 2011, yielded a sample of 8,245 persons with a 71% weighted response rate. Proxy respondents were interviewed when the participant could not respond (n=583 or 5.8% [weighted]).^25,26^ The protocol was approved by the Johns Hopkins Institutional Review Board, and all participants provided informed consent.

CMS records of fee-for-service Medicare claims, cross-linked to NHATS data, were used to identify participants who underwent major surgery. Comparable data are not available in NHATS from Medicare Advantage. Major surgery was defined as any procedure in an operating room requiring the use of general anesthesia for a non-percutaneous, non-endoscopic, invasive operation. This definition, which has been previously implemented by our group,^12,27^ is consistent with other definitions of “high-risk” surgery in older persons.^28,29^ We categorized each procedure into one of six subtypes: 1. Musculoskeletal; 2. Abdominal/Gastrointestinal; 3. Vascular (including endovascular, non-coronary bypass grafts, and amputations); 4. Neurologic (including brain and spine); 5. Cardiothoracic; and 6. Other (including major endocrine, gynecologic, urologic, breast, plastic, otolaryngologic, and transplant surgery). Major surgeries were categorized as elective (planned) or non-elective (unplanned) based on a CMS indicator variable.^12,27,30^ Ophthalmology procedures did not meet criteria for major surgery.

### Study Population

Among the 7,609 NHATS participants who were living in settings other than nursing homes (community-living) at the time of their Round 1 interview, we identified those who were enrolled for at least 1 month in fee-for-service Medicare during the subsequent 5-year surveillance window from 2011 to 2016. The number of participants with continuous fee-for-service Medicare, a combination of fee-for-service Medicare and Medicare Advantage, and Medicare Advantage only were 4,559 (61.1%), 1,012 (13.1%), and 2,038 (25.8%), respectively.

During Round 1 of NHATS, information was collected on demographic characteristics, including age, sex, race/ethnicity, education level, whether the individual lives alone; ten self-reported, physician diagnosed chronic conditions, including heart attack, high blood pressure, arthritis, osteoporosis, diabetes, lung disease, stroke, dementia or Alzheimer’s disease, cancer, hip fracture since age 50; and two geriatric characteristics: frailty and dementia.^26^ Participants were categorized as non-frail, pre-frail, and frail according to the Fried phenotype^31^ and as having no dementia, possible dementia, or probable dementia based on a validated assessment strategy.^32,33^ Data on frailty and dementia were 100% complete.^31,32^ Medicaid eligibility was obtained from CMS data.

### Statistical Analysis

#### Incidence Rates

We calculated nationally-representative incidence rates of major surgery over the 5-year follow-up period using Poisson regression. All eligible surgeries were included in the numerator of a given rate; to account for differences in time at risk for surgery, we used participants’ time (in months) with fee-for-service Medicare coverage as the offset term. We calculated overall (unadjusted) incidence rates per 100 person-years for all major surgeries and separately by timing (elective and non-elective) and subtype of surgery. In addition, for all major surgeries, elective surgeries, and non-elective surgeries, we calculated both unadjusted and age-and sex-adjusted incidence rates stratified by four demographic and geriatric characteristics: age, sex, frailty, and dementia.

#### Cumulative Risk

We calculated the proportion (cumulative risk) and incidence count of individuals who underwent at least one major surgery during the 5-year follow-up period using data from NHATS participants who were continuously enrolled in fee-for-service Medicare, including decedents.^34,35^ Estimates were calculated for all major surgery, elective and non-elective major surgery, and subgroups defined by age, sex, frailty, and dementia.

To generate nationally-representative incidence and cumulative risk estimates, all analyses incorporated both the NHATS Round 1 analytic sampling weights^25,36,37^ and the cluster and strata variables, thereby accounting for the complex sample survey design.^25^ All analyses were performed using SAS version 9.4 (SAS Institute, Inc., Cary, North Carolina, US). This study followed the Strengthening the Reporting of Observational Studies in Epidemiology (STROBE) reporting guideline.^38^

## RESULTS

Among the 5,571 community-living NHATS participants who were enrolled in fee-for-service Medicare for at least 1 month between May 2011 and November 2016 (**Table 1**), the mean age was 75.3 (Standard Error, 0.1) years, more than half were female (55.9%), and 4 out of 5 (81.5%) were non-Hispanic White. More than 1 in 8 (13.4%) were Medicaid eligible, and nearly 1 in 2 (43.4%) had 3 or more chronic conditions. Almost half (46.7%) of the participants were pre-frail, and 17.2% were frail, while more than 1 in 5 were cognitively impaired, with either possible (10.7%) or probable (10.7%) dementia. Differences between NHATS participants with at least 1 month of fee-for-service Medicare and those in fee-for-service only or Medicare Advantage only were relatively small, with the latter group having a slightly lower percentage of males, living alone, Medicaid eligible, frailty, and probable dementia.

**Table 1:** Characteristics of Community-Living NHATS Participants (N=7,609) According to Type of Medicare Coverage During 5-year Follow-up Period, 2011 - 2016^a^.

| Variable | At Least 1 Month Fee-for-Service Coverage (N=5571) |  | Fee-for-Service Only (N=4559) |  | Mixed Fee-for-Service and Medicare Advantage (N=1012) |  | Medicare Advantage Only (N=2038) |  |
| --- | --- | --- | --- | --- | --- | --- | --- | --- |
|  | N | % or mean (SE) | N | % or mean (SE) | N | % or mean (SE) | N | % or mean (SE) |
| Mean age, years | 5571 | 75.3 (0.10) | 4559 | 75.7 (0.11) | 1012 | 73.7 (0.22) | 2038 | 75.2 (0.16) |
| Age groups (in years) |  |  |  |  |  |  |  |  |
| 65-69 | 1049 | 28.4 | 805 | 27.2 | 244 | 34.1 | 360 | 26.6 |
| 70-74 | 1114 | 24.3 | 870 | 23.6 | 244 | 27.7 | 465 | 26.8 |
| 75-79 | 1073 | 18.6 | 870 | 18.6 | 203 | 18.3 | 440 | 20.5 |
| 80-84 | 1105 | 14.7 | 951 | 15.7 | 154 | 10.3 | 400 | 14.6 |
| 85-89 | 737 | 9.6 | 625 | 10.0 | 112 | 7.3 | 216 | 7.8 |
| 90 and older | 493 | 4.4 | 438 | 4.9 | 55 | 2.4 | 157 | 3.7 |
| Gender |  |  |  |  |  |  |  |  |
| Male | 2362 | 44.1 | 1950 | 44.1 | 412 | 44.0 | 809 | 41.5 |
| Female | 3209 | 55.9 | 2609 | 55.9 | 600 | 56.0 | 1229 | 58.6 |
| Race & Ethnicity |  |  |  |  |  |  |  |  |
| White, non-Hispanic | 3851 | 81.5 | 3320 | 84.1 | 531 | 69.3 | 1335 | 77.7 |
| Black, non-Hispanic | 1192 | 7.8 | 855 | 6.7 | 337 | 13.0 | 470 | 9.0 |
| Hispanic | 286 | 5.7 | 202 | 4.7 | 84 | 10.4 | 168 | 9.7 |
| Other | 173 | 3.7 | 124 | 3.2 | 49 | 6.2 | 46 | 2.8 |
| Unknown | 69 | 1.3 | 58 | 1.3 | 11 | 1.2 | 19 | 0.8 |
| Education |  |  |  |  |  |  |  |  |
| < High school | 1485 | 20.9 | 1136 | 19.6 | 349 | 27.1 | 562 | 23.1 |
| High school or equivalent | 1460 | 26.5 | 1217 | 26.8 | 243 | 24.9 | 609 | 29.5 |
| > High School | 2554 | 51.3 | 2148 | 52.3 | 406 | 46.4 | 843 | 46.3 |
| Unknown | 72 | 1.4 | 58 | 1.3 | 14 | 1.6 | 24 | 1.0 |

(Table 1 continued)
| Variable | At Least 1 Month Fee-for-Service Coverage (N=5571) |  | Fee-for-Service Only (N=4559) |  | Mixed Fee-for-Service and Medicare Advantage (N=1012) |  | Medicare Advantage Only (N=2038) |  |
| --- | --- | --- | --- | --- | --- | --- | --- | --- |
|  | N | % or mean (SE) | N | % or mean (SE) | N | % or mean (SE) | N | % or mean (SE) |
| Lives alone | 1870 | 30.6 | 1522 | 30.7 | 348 | 30.0 | 618 | 27.6 |
| Medicaid eligible | 899 | 13.4 | 626 | 11.2 | 273 | 23.4 | 265 | 10.4 |
| Number of chronic conditions (0-10) | 5570 | 2.4 (0.02) | 4558 | 2.4 (0.02) | 1012 | 2.3 (0.05) | 2038 | 2.3 (0.03) |
| 3 or more | 2605 | 43.4 | 2158 | 44.0 | 447 | 40.4 | 938 | 43.2 |
| Frail phenotype |  |  |  |  |  |  |  |  |
| Non-frail | 1703 | 36.1 | 1403 | 36.5 | 300 | 34.4 | 674 | 38.4 |
| Pre-frail | 2660 | 46.7 | 2178 | 46.6 | 482 | 47.3 | 957 | 45.4 |
| Frail | 1208 | 17.2 | 978 | 16.9 | 230 | 18.3 | 407 | 16.2 |
| Dementia status |  |  |  |  |  |  |  |  |
| No dementia | 4036 | 78.7 | 3315 | 78.9 | 721 | 77.5 | 1539 | 80.1 |
| Possible dementia | 721 | 10.7 | 574 | 10.6 | 147 | 11.2 | 275 | 11.6 |
| Probable dementia | 814 | 10.7 | 670 | 10.5 | 144 | 11.3 | 224 | 8.3 |

Abbreviations: NHATS, National Health and Aging Trends Study; N, number; SE, standard error.
<sup>a</sup> N's are unweighted; means, percentages and standard errors are weighted.

### Incidence Rates

Based on a total estimated time at risk of 97,049,615 person-years, the nationally-representative incidence rate of major surgery was 8.8 per 100 person-years (95% confidence interval [CI] 8.2-9.5), indicating that for every 100 community-living older Americans, 8.8 major surgeries were performed annually in the US. The national rates of elective and non-elective surgery were 5.2 (95% CI, 4.7-5.7) and 3.7 (95% CI, 3.3-4.1), respectively.

The adjusted incidence rate per 100 person-years of all major surgeries varied by age group (**Figure 1**), peaking at 10.8 (95% CI, 9.4-12.4) in persons 75-79 years and declining to a low of 6.4 (95% CI, 5.0-8.2) in those ≥90 years. Rates of elective surgery were highest in the youngest three age groups (65-69; 70-74; 75-79), while rates of non-elective surgery were highest in the oldest three age groups (80-84; 85-89; ≥90). Rates were similar between males and females for both elective and non-elective surgeries. The incidence of major surgery increased with worsening frailty, from 6.6 (95% CI, 5.8-7.5) in the non-frail group to 10.3 (95% CI, 8.9-11.9) in the frail group. This increase was driven largely by an increase in non-elective surgery with worsening frailty, from 2.6 (95% CI, 2.1-3.2) in the non-frail group to 5.5 (95% CI, 4.5-6.6) in the frail group. Although there was relatively little difference in the rate of major surgery across the three dementia groups, the rate of elective surgery was considerably higher in the no dementia (4.2 [95% CI, 3.7-4.7]) than the probable dementia group (2.4 [95% CI, 1.6-3.6]), while the rate of non-elective surgery was considerably higher in the probable dementia (5.3 [95% CI, 4.2-6.7)]) than no dementia group (3.9 [95% CI, 3.4-4.4]). The unadjusted and adjusted rates by age, sex, frailty, and dementia are shown in **eTable 1**, which also provides unadjusted rates by subtype of surgery. The most frequent per 100 person-years were musculoskeletal (3.6 [95% CI, 3.2-4.0]) and abdominal/gastrointestinal (1.7 [95% CI, 1.4-2.0]), while the least frequent was cardiothoracic (0.7 [95% CI, 0.6-0.9]).

**FIGURE 1.**
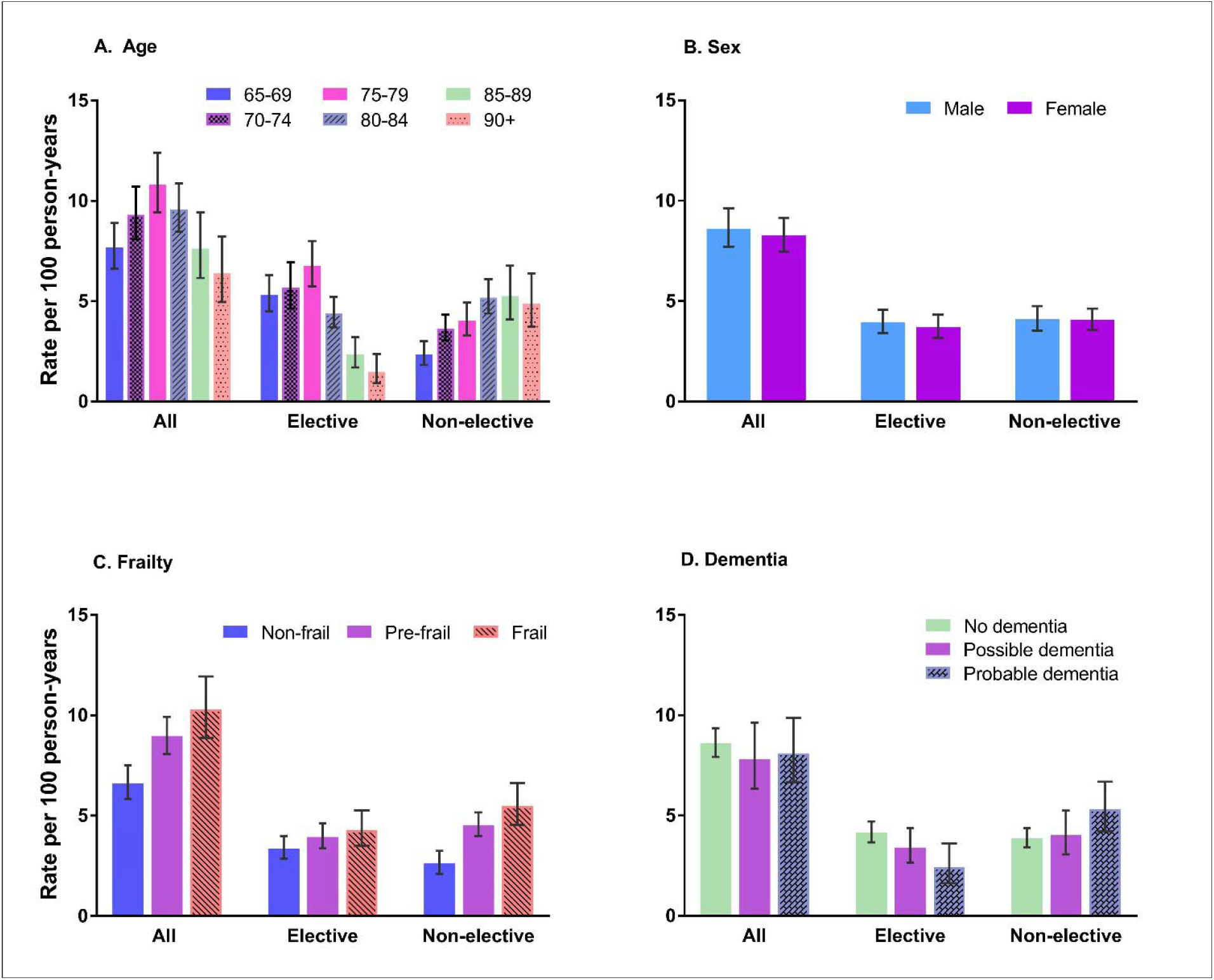
Age- and sex-adjusted incidence rates per 100 person-years of major surgery by age (A), sex (B), frailty (C), and dementia (D). All plots are shown by timing of surgery (elective vs non-elective). Major surgery subtypes are aggregated. Error bars represent 95% confidence intervals. Exact incidence rates are provided in **eTable 1**.

### Cumulative Risk

The 5-year cumulative risk of major surgery, based on participants who were continuously enrolled in fee-for-service Medicare, was 13.8% (95% CI, 12.2%-15.5%), representing nearly 5 million unique older persons (4,958,048 [95% CI, 4,345,342-5,570,755]). The cumulative risks were 8.6% (95% CI, 7.3%-9.2%) for elective surgery, yielding nearly 3.1 million older persons (3,088,809; [95% CI, 2,600,831-3,576,187]) and 6.8% (95% CI, 6.0%-7.6%) for non-elective surgery, yielding nearly 2.5 million older persons (2,439,253; [95% CI, 2,157,235-2,721,272]).

**Figure 2** shows the 5-year cumulative risk of major surgery by the four demographic and geriatric characteristics. Values ranged from more than 9% in persons ≥90 years to more than 16% in the 74-79 and 80-84 age groups. The cumulative risk was not evenly distributed between elective and non-elective surgeries by age: the younger age groups (65-69; 70-74; 75-79) had higher risk for elective surgery, while the older age groups (80-84; 85-89; ≥90) had higher risk for non-elective surgery. The cumulative risk of major surgery was nearly identical for males and females, for both elective and non-elective surgeries. Values for cumulative risk were 15.6% (95% CI, 13.6%-17.5%) and 12.1% (95% CI, 9.9%-14.4%) in the pre-frail and frail groups. For both groups, risk was about equally divided between elective and non-elective major surgery, although the frail group had a slightly higher cumulative risk of non-elective surgery. In contrast, the cumulative risk of elective surgery (9.2% [95% CI, 7.3%-11.1%]) was much greater than that of non-elective surgery (4.5% [95% CI, 3.5%-5.5%]) in the non-frail group, yielding an overall risk of 12.6% (95% CI, 10.6%-14.5%). Although the cumulative risk of all major surgeries differed little by dementia status, the risk of elective surgery was highest in the no dementia group (9.4% [95% CI, 7.9%-10.8%]), while the risk of non-elective surgery was highest in the dementia group (8.9% [95% CI, 6.5%-11.4%]).

**FIGURE 2:**
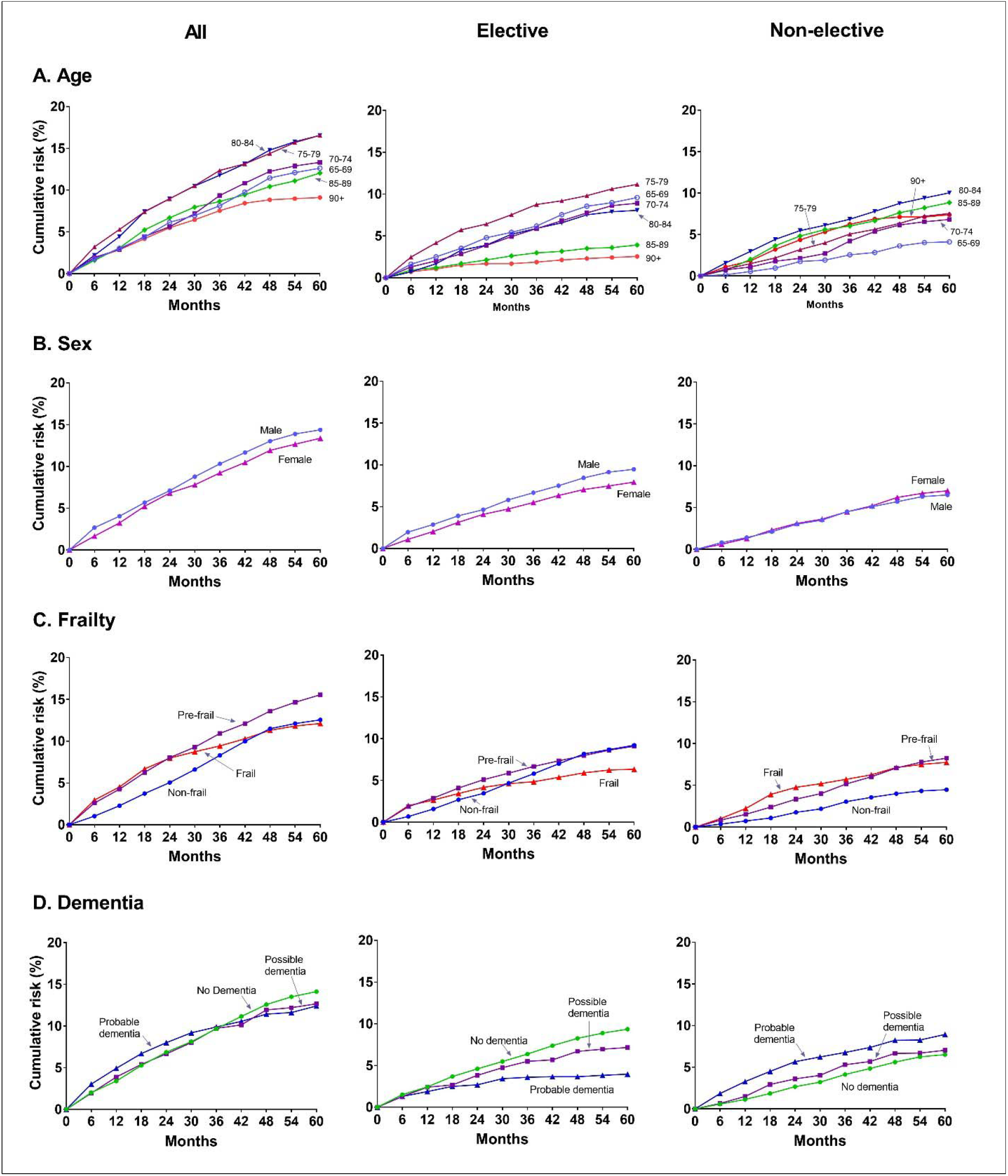
Cumulative risk of any major surgery over 5 years by age (A), sex (B), frailty (C), and dementia (D). Plots are shown by all major surgery as well as elective and non-elective major surgery. Major surgery subtypes are aggregated. Exact weighted cumulative risk proportions, with 95% confidence intervals, are provided in **eTable 2**.

More detailed information about the cumulative risk of major surgery is provided in **eTable 2**.

## DISCUSSION

In this nationally-representative sample of community-living older persons, we estimated the incidence and cumulative risk of major surgery in the US over a 5-year period and evaluated how these estimates differ by demographic and geriatric characteristics for both elective and non-elective surgeries. Our findings demonstrate that major surgery is a common event in the lives of community-living older persons, including high-risk vulnerable subgroups such as the oldest old (≥85 years), those with frailty or dementia, and those undergoing non-elective surgery. As the US prepares to meet the challenges of an aging society,^2,39^ the burden of major surgery in older Americans warrants attention from policy makers and others committed to a high-functioning national health care system.

We found that nearly 9 major surgeries were performed annually for every 100 community-living persons aged 65 years or older. During the 5-year surveillance window, more than 1 in 7 Medicare beneficiaries underwent at least one major surgery, representing nearly 5 million unique older persons. Two in 5 of these major surgeries were unplanned. Vulnerable subgroups had amongst the highest incidence rates and cumulative risks of major surgery in general and non-elective surgery in particular.

Our findings have important implications for the US health care system in terms of costs, caregivers, and care. First, the growing number of Medicare beneficiaries requiring major surgery in the coming years will further stress an already expensive Medicare program. In 2018, Medicare spending was $605 billion, representing 20% of total national health spending, 15% of the entire federal budget, and 3% of gross domestic product (GDP).^40^ During the next 30 years, with an increasing number of beneficiaries, Medicare spending is projected to grow considerably – to 6% of GDP.^41^ As over half of fee-for-service Medicare expenditures are for surgical care, and nearly 50% of hospitalization costs are related to operating room-based procedures,^42,43^ the ability of Medicare to continue to cover all major surgery in the future will be fiscally challenging.

Second, there will be increased demand for surgeons and a perioperative workforce with competency in geriatric care. The US is already experiencing shortages in nine of 10 surgical specialties, with orthopedic surgery, general surgery, and urology having the largest surgeon deficits for specialties performing major surgery.^44,45^ Based on current projections suggesting that the supply of surgeons will remain constant for the next 15 years, the US will lack as many as 28,700 surgeons by 2033.^46^ Furthermore, as highlighted in an Institute of Medicine report, the elder care workforce to manage geriatric patients is inadequate.^47^ Given the multidisciplinary nature of perioperative management, the predicted shortfall of up to 139,000 physicians and 918,000 registered nurses within the next 10-13 years may adversely affect the ability to perform timely, safe, and high-quality major surgery.^46,48^

Third, innovative strategies will be required to better address and prioritize the unique needs and wishes of older surgical patients in the perioperative period, especially those in high-risk vulnerable subgroups. Major surgery is inherently invasive and may be unnecessary, harmful, and even futile in some older persons. Nearly one-third of fee-for-service Medicare beneficiaries have at least one major surgery in the year before death, including 20% in the last month of life.^29^ While studies of decedents must be interpreted carefully,^49^ a large proportion of major surgeries are performed near the end of life. When surgery is being considered, the outcomes that matter most to older persons are symptom burden, functional independence, and health-related quality of life.^50–52^ To reduce non-patient-centered, non-beneficial surgical care,^53–57^ developing new approaches to perioperative decision making, risk stratification, outcome metrics, and goals of care will be needed. Future efforts should build on ongoing work by the Coalition for Quality in Geriatric Surgery (CQGS), a partnership between the American College of Surgeons (ACS) and the John A. Hartford Foundation.^17^

Measuring the current and future population-based burden of conditions that have a large-scale impact on public health is a fundamental feature of a mature, high-performance health care system.^58–62^ Despite its importance to public health and policy, up-to-date and reliable data on the occurrence of major surgery in the US are not readily available. The present analysis addresses this knowledge gap by providing current estimates on the incidence and cumulative risk of major surgery in older persons. These estimates will strengthen and enhance policymaking, allow health care institutions to more accurately plan for the future, and reinforce key areas of clinical practice that need to be addressed.

Three unique strengths enhance the generalizability, validity, and applicability of our findings. First, by linking CMS data to NHATS, a population-based cohort, we were able to generate nationally-representative estimates of major surgery in Medicare beneficiaries for the contiguous US. These estimates included both incidence and cumulative risk over a 5-year period, two complementary metrics of utilization. Prior studies employed only regional or state-level assessments, which do not provide a comprehensive picture of major surgery in the US.^18^ Other studies are outdated, often by more than 20 years, and are no longer relevant.^4,18–20,28^ Second, we use an established definition of major surgery in older persons that is clearly defined, clinically relevant, widely accepted, and encompasses the spectrum of surgical disciplines.^12,27–29^ Prior studies that have estimated rates of geriatric surgery employed either an overly-broad definition of a procedure,^4,19–21,63^ including very minor diagnostic and therapeutic interventions, or a very narrow definition, including only a small handful of operations.^18^ These definitions could lead to erroneous conclusions about the rates and risks of major surgery in older persons and contribute to misleading population-based estimates. Third, we provide estimates for subgroups defined on the basis of frailty and dementia, two key determinants of health and well-being in older persons.^12,27,64–66^ To our knowledge, prior studies that have estimated rates of major surgery in older persons have not included geriatric characteristics,^5,18–23^ thereby limiting their applicability.

Our findings should be interpreted in the context of the following limitations. First, our results are limited to fee-for-service Medicare beneficiaries since CMS data on Medicare Advantage were not available. The penetrance of Medicare Advantage was around 25% in the current study but is projected to increase to 42% by 2028.^67^ With the recent decision by CMS to make Medicare Advantage claims data more broadly available,^68^ it should be possible to base future estimates on all Medicare beneficiaries. Second, because information on the geriatric conditions was not updated beyond NHATS Round 1, our findings may underestimate the occurrence of major surgeries in older persons who are frail or have dementia since the prevalence of these conditions increases over time. Third, we focused solely on the incidence and cumulative risk of major surgery in older persons. Future studies using nationally-representative data sources of older persons should focus on other high priority areas, such as evaluating functional outcomes, mortality, and quality of life after major surgery and elucidating potential health disparities related to major surgery, from racial and ethnic to geographic and socioeconomic.

## CONCLUSION

Major surgery is a defining health issue for community-living older persons, including high-risk vulnerable subgroups such as the oldest old, those with frailty or dementia, and those having non-elective major surgery. Our findings provide up-to-date, generalizable data on the occurrence of major surgery in older Americans and highlight potential challenges for the US health care system in the context of an aging population needing major surgery.

## Data Availability

Because of DUA restrictions, we cannot provide external investigators with access to analysis-specific NHATS cohort study data that include sensitive/restricted files. Upon request, we will make the code that constructed files and/or ran analyses available to external investigators who wish to replicate our analyses.

## Authors’ Contributions

*Concept and design*: Becher, Gill

*Acquisition, analysis, or interpretation of the data*: All authors

*Drafting of the manuscript*: All authors

*Critical revision of the manuscript for important intellectual content*: All authors

*Statistical Analysis*: Becher, Vander Wyk, Leo-Summers, Gill

*Administrative, technical, or material support*: Becher, Vander Wyk, Leo-Summers, Gill

*Supervision*: Becher, Gill

## Data Access, Responsibility, and Analysis

Becher, Vander Wyk, and Gill had full access to all the data in the study and take responsibility for the integrity of the data and the accuracy of the data analysis.

## Supplementary Online Content

**eTable 1:**
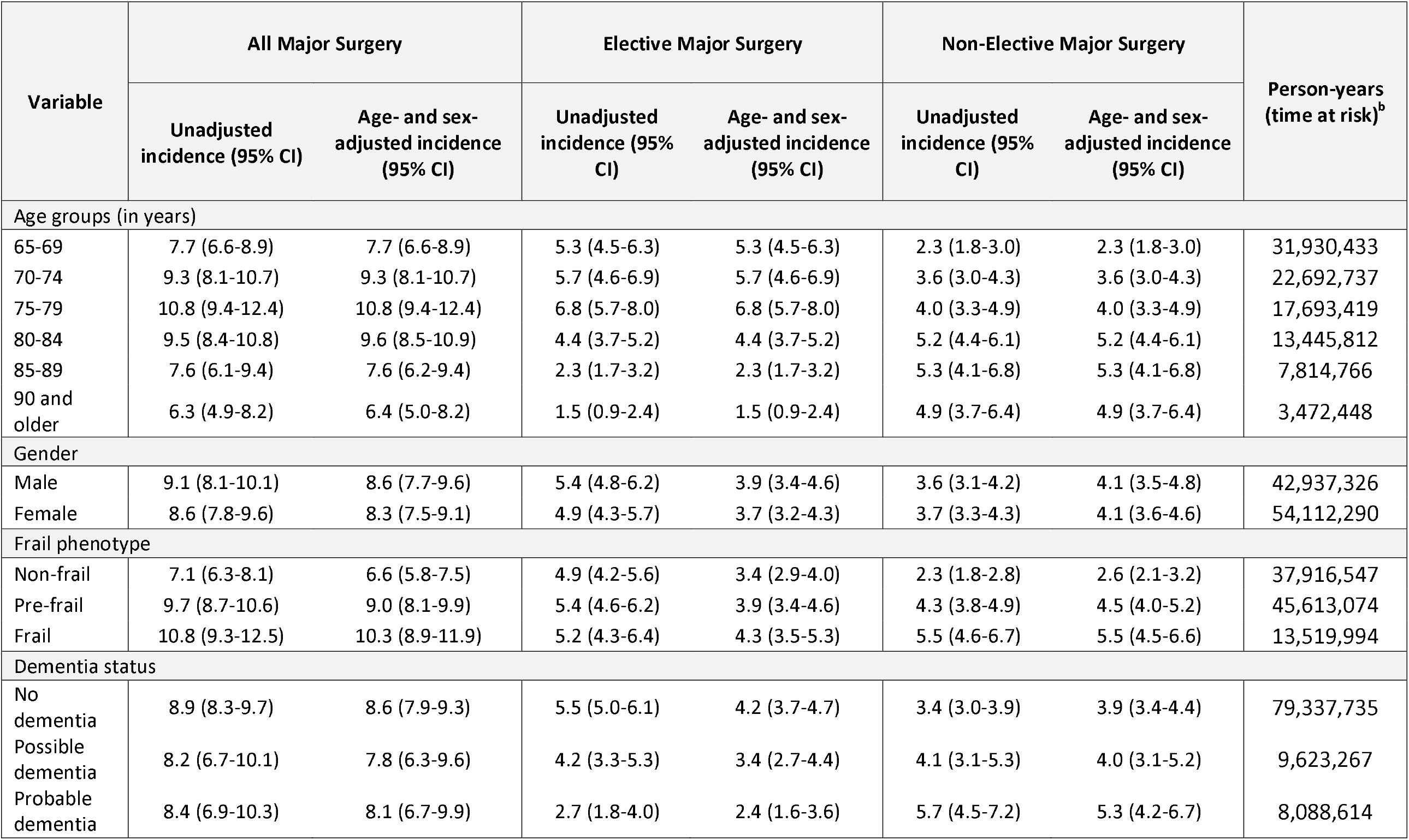
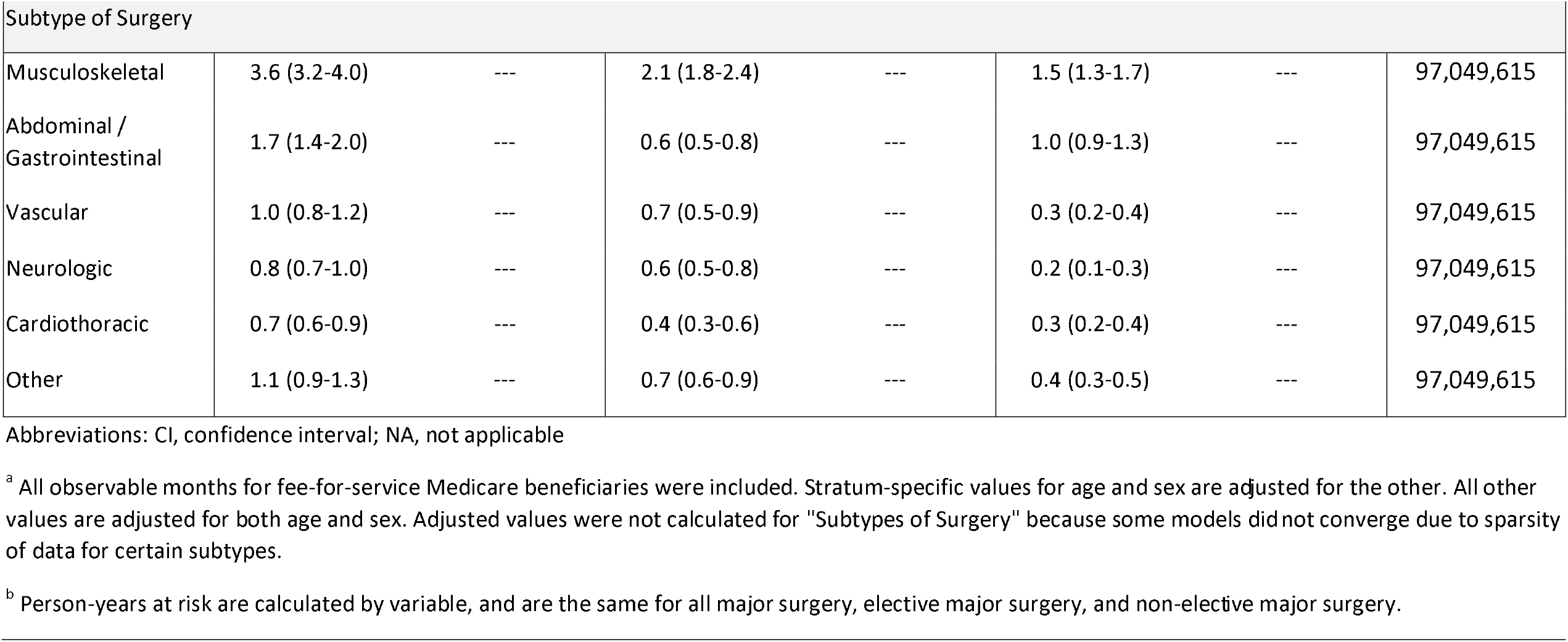
Incidence Rates of Major Surgery in Medicare Beneficiaries According to Demographic and Geriatric Characteristics and Subtype of Surgery, by Timing of Operation^a^.

| Variable | All Major Surgery |  | Elective Major Surgery |  | Non-Elective Major Surgery |  | Person-years<br>(time at risk)<sup>b</sup> |
| --- | --- | --- | --- | --- | --- | --- | --- |
|  | Unadjusted<br>incidence (95% CI) | Age- and sex-<br>adjusted incidence<br>(95% CI) | Unadjusted<br>incidence (95%<br>CI) | Age- and sex-<br>adjusted incidence<br>(95% CI) | Unadjusted<br>incidence (95%<br>CI) | Age- and sex-<br>adjusted incidence<br>(95% CI) |  |
| Age groups (in years) |  |  |  |  |  |  |  |
| 65-69 | 7.7 (6.6-8.9) | 7.7 (6.6-8.9) | 5.3 (4.5-6.3) | 5.3 (4.5-6.3) | 2.3 (1.8-3.0) | 2.3 (1.8-3.0) | 31,930,433 |
| 70-74 | 9.3 (8.1-10.7) | 9.3 (8.1-10.7) | 5.7 (4.6-6.9) | 5.7 (4.6-6.9) | 3.6 (3.0-4.3) | 3.6 (3.0-4.3) | 22,692,737 |
| 75-79 | 10.8 (9.4-12.4) | 10.8 (9.4-12.4) | 6.8 (5.7-8.0) | 6.8 (5.7-8.0) | 4.0 (3.3-4.9) | 4.0 (3.3-4.9) | 17,693,419 |
| 80-84 | 9.5 (8.4-10.8) | 9.6 (8.5-10.9) | 4.4 (3.7-5.2) | 4.4 (3.7-5.2) | 5.2 (4.4-6.1) | 5.2 (4.4-6.1) | 13,445,812 |
| 85-89 | 7.6 (6.1-9.4) | 7.6 (6.2-9.4) | 2.3 (1.7-3.2) | 2.3 (1.7-3.2) | 5.3 (4.1-6.8) | 5.3 (4.1-6.8) | 7,814,766 |
| 90 and<br>older | 6.3 (4.9-8.2) | 6.4 (5.0-8.2) | 1.5 (0.9-2.4) | 1.5 (0.9-2.4) | 4.9 (3.7-6.4) | 4.9 (3.7-6.4) | 3,472,448 |
| Gender |  |  |  |  |  |  |  |
| Male | 9.1 (8.1-10.1) | 8.6 (7.7-9.6) | 5.4 (4.8-6.2) | 3.9 (3.4-4.6) | 3.6 (3.1-4.2) | 4.1 (3.5-4.8) | 42,937,326 |
| Female | 8.6 (7.8-9.6) | 8.3 (7.5-9.1) | 4.9 (4.3-5.7) | 3.7 (3.2-4.3) | 3.7 (3.3-4.3) | 4.1 (3.6-4.6) | 54,112,290 |
| Frail phenotype |  |  |  |  |  |  |  |
| Non-frail | 7.1 (6.3-8.1) | 6.6 (5.8-7.5) | 4.9 (4.2-5.6) | 3.4 (2.9-4.0) | 2.3 (1.8-2.8) | 2.6 (2.1-3.2) | 37,916,547 |
| Pre-frail | 9.7 (8.7-10.6) | 9.0 (8.1-9.9) | 5.4 (4.6-6.2) | 3.9 (3.4-4.6) | 4.3 (3.8-4.9) | 4.5 (4.0-5.2) | 45,613,074 |
| Frail | 10.8 (9.3-12.5) | 10.3 (8.9-11.9) | 5.2 (4.3-6.4) | 4.3 (3.5-5.3) | 5.5 (4.6-6.7) | 5.5 (4.5-6.6) | 13,519,994 |
| Dementia status |  |  |  |  |  |  |  |
| No<br>dementia | 8.9 (8.3-9.7) | 8.6 (7.9-9.3) | 5.5 (5.0-6.1) | 4.2 (3.7-4.7) | 3.4 (3.0-3.9) | 3.9 (3.4-4.4) | 79,337,735 |
| Possible<br>dementia | 8.2 (6.7-10.1) | 7.8 (6.3-9.6) | 4.2 (3.3-5.3) | 3.4 (2.7-4.4) | 4.1 (3.1-5.3) | 4.0 (3.1-5.2) | 9,623,267 |
| Probable<br>dementia | 8.4 (6.9-10.3) | 8.1 (6.7-9.9) | 2.7 (1.8-4.0) | 2.4 (1.6-3.6) | 5.7 (4.5-7.2) | 5.3 (4.2-6.7) | 8,088,614 |
|  | Unadjusted<br>incidence (95% CI) | Age- and sex-<br>adjusted incidence<br>(95% CI) | Unadjusted<br>incidence (95% CI) | Age- and sex-<br>adjusted incidence<br>(95% CI) | Unadjusted<br>incidence (95% CI) | Age- and sex-<br>adjusted incidence<br>(95% CI) |  |
| Subtype of Surgery |  |  |  |  |  |  |  |
| Musculoskeletal | 3.6 (3.2-4.0) | --- | 2.1 (1.8-2.4) | --- | 1.5 (1.3-1.7) | --- | 97,049,615 |
| Abdominal /<br>Gastrointestinal | 1.7 (1.4-2.0) | --- | 0.6 (0.5-0.8) | --- | 1.0 (0.9-1.3) | --- | 97,049,615 |
| Vascular | 1.0 (0.8-1.2) | --- | 0.7 (0.5-0.9) | --- | 0.3 (0.2-0.4) | --- | 97,049,615 |
| Neurologic | 0.8 (0.7-1.0) | --- | 0.6 (0.5-0.8) | --- | 0.2 (0.1-0.3) | --- | 97,049,615 |
| Cardiothoracic | 0.7 (0.6-0.9) | --- | 0.4 (0.3-0.6) | --- | 0.3 (0.2-0.4) | --- | 97,049,615 |
| Other | 1.1 (0.9-1.3) | --- | 0.7 (0.6-0.9) | --- | 0.4 (0.3-0.5) | --- | 97,049,615 |

Abbreviations: CI, confidence interval; NA, not applicable
<sup>a</sup> All observable months for fee-for-service Medicare beneficiaries were included. Stratum-specific values for age and sex are adjusted for the other. All other values are adjusted for both age and sex. Adjusted values were not calculated for "Subtypes of Surgery" because some models did not converge due to sparsity of data for certain subtypes.
<sup>b</sup> Person-years at risk are calculated by variable, and are the same for all major surgery, elective major surgery, and non-elective major surgery.

**eTable 2:**
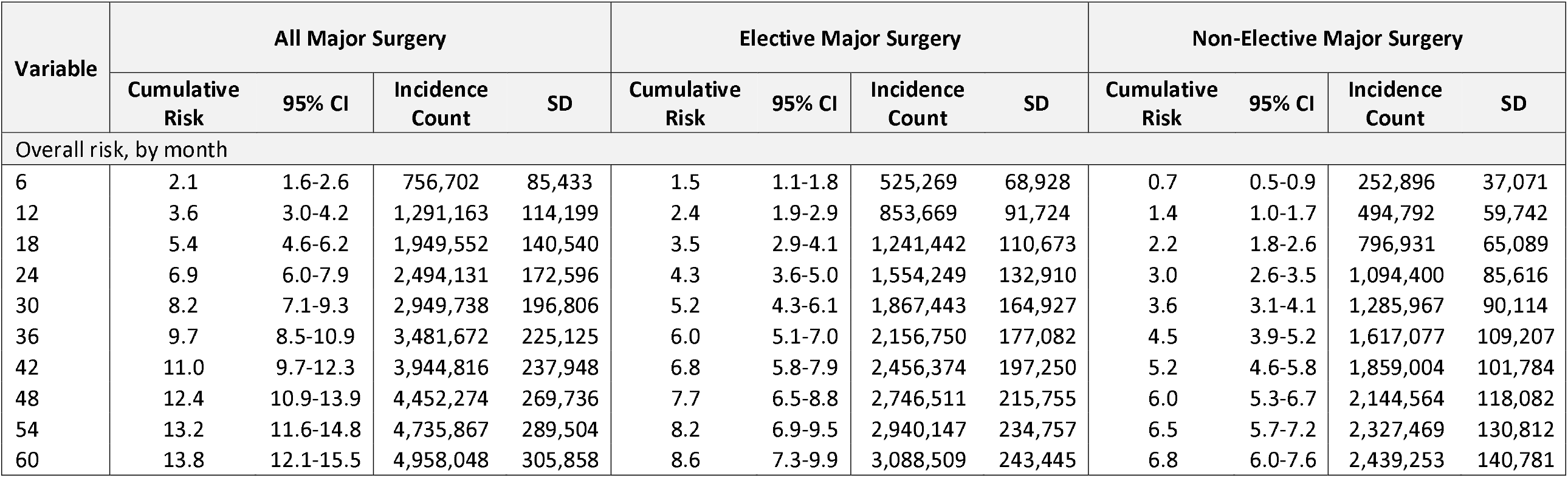
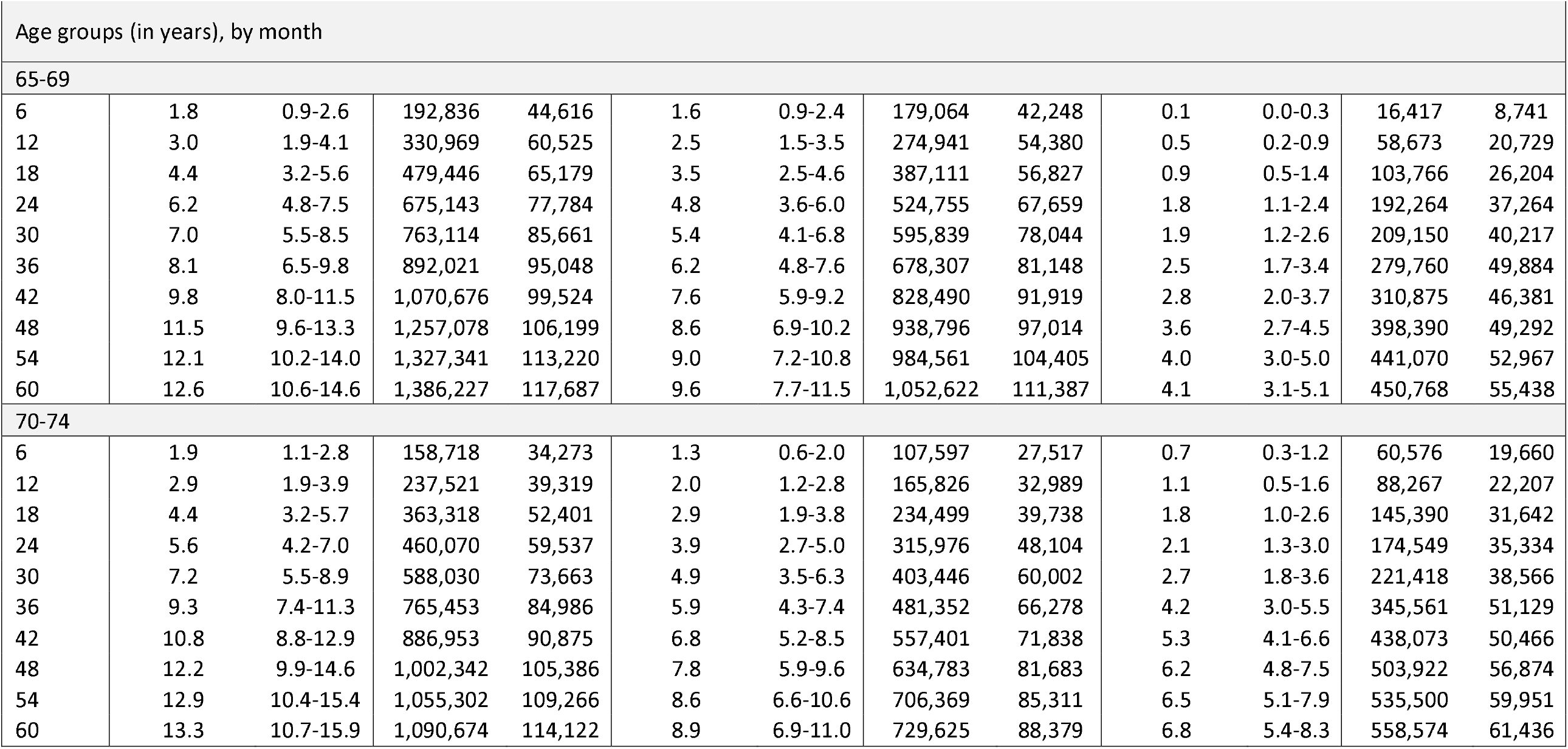
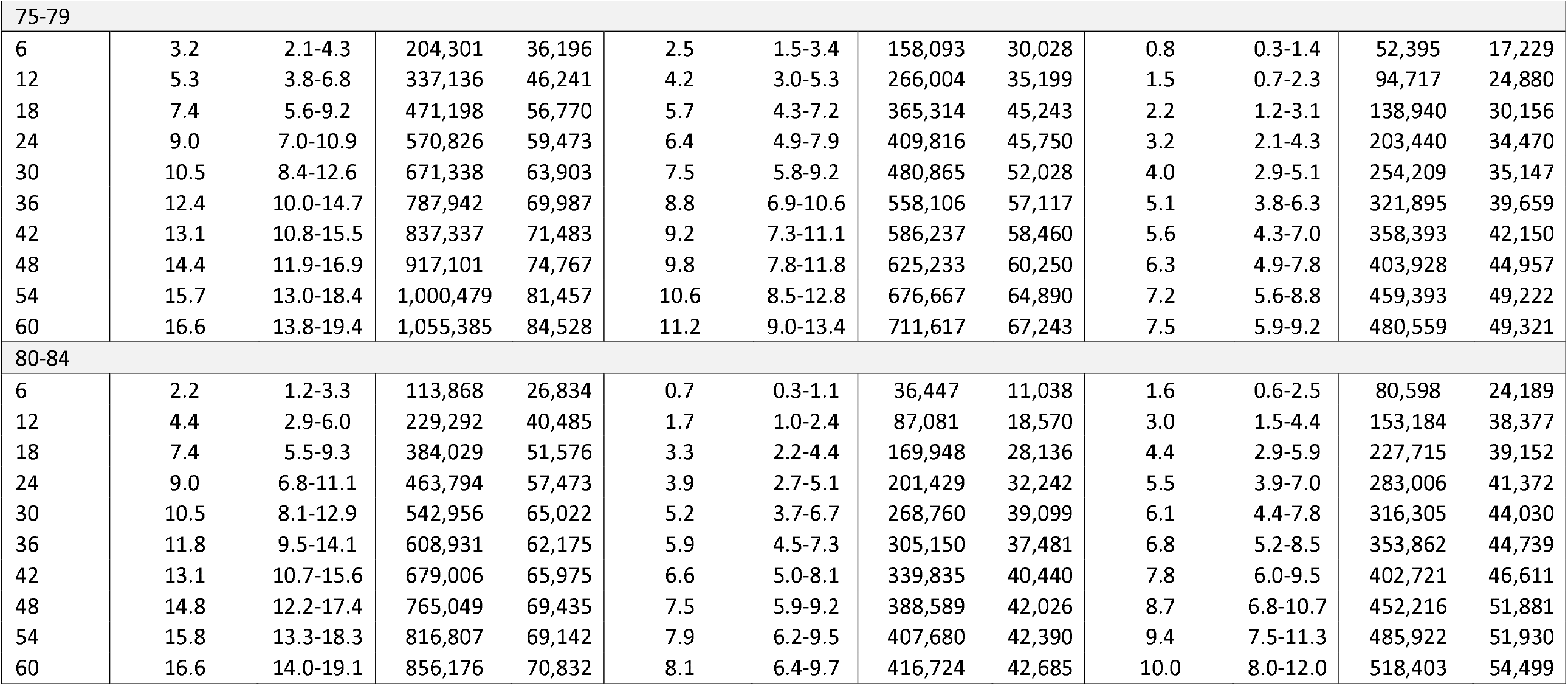
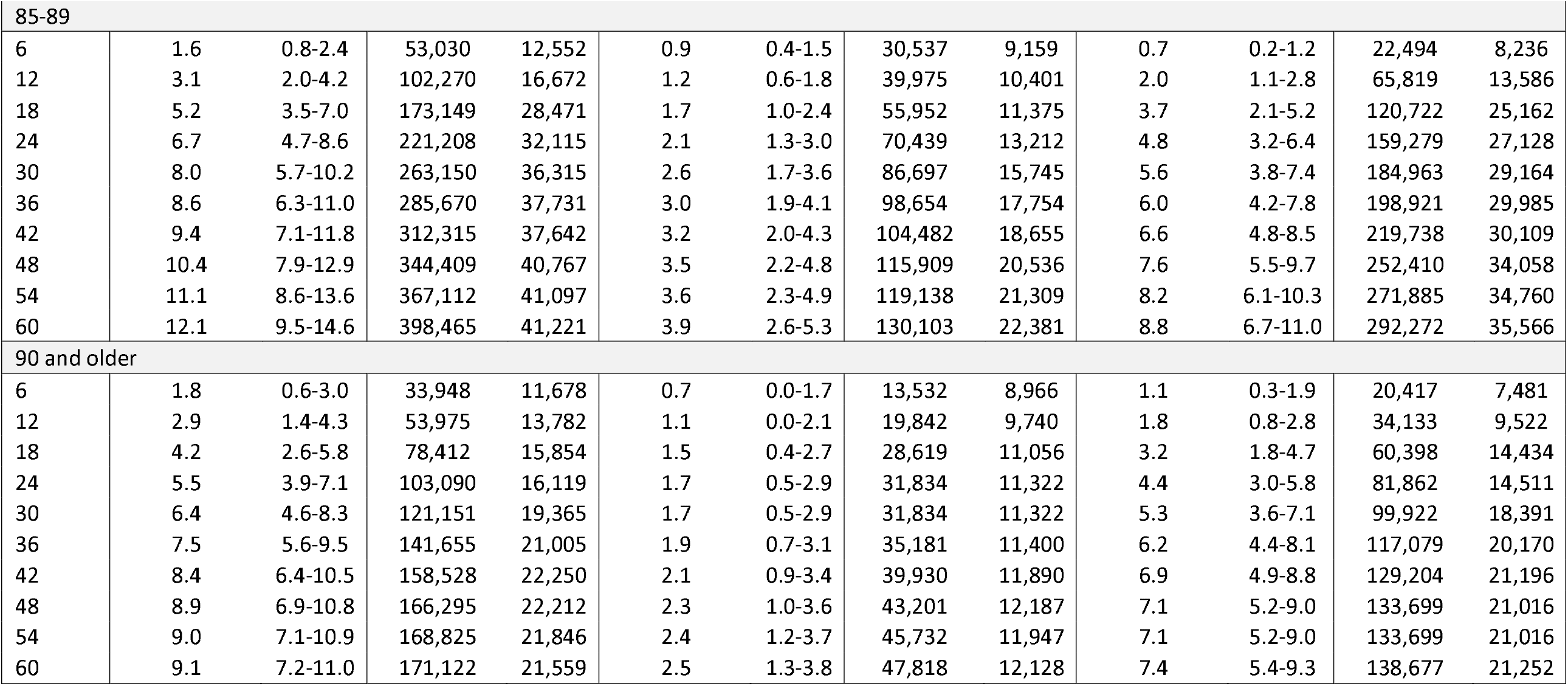
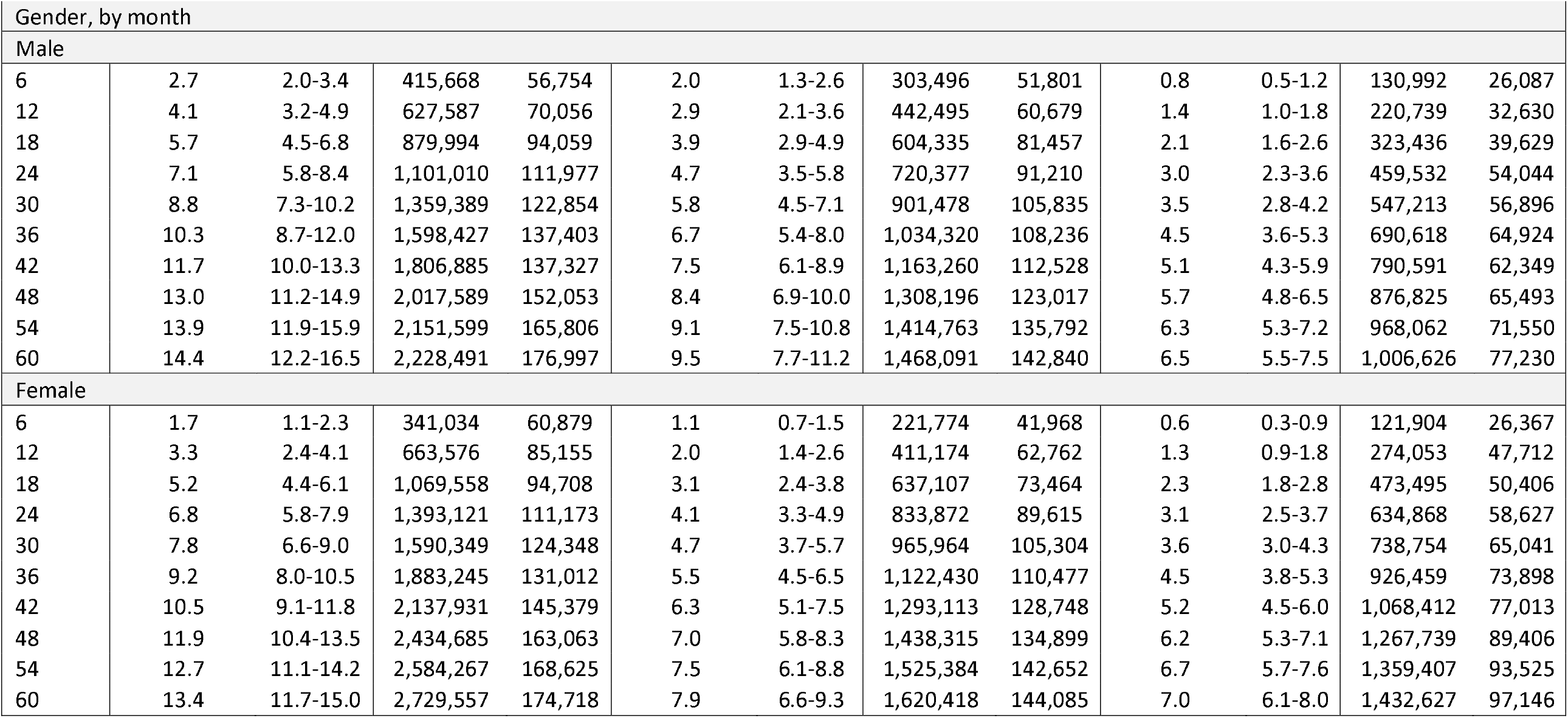
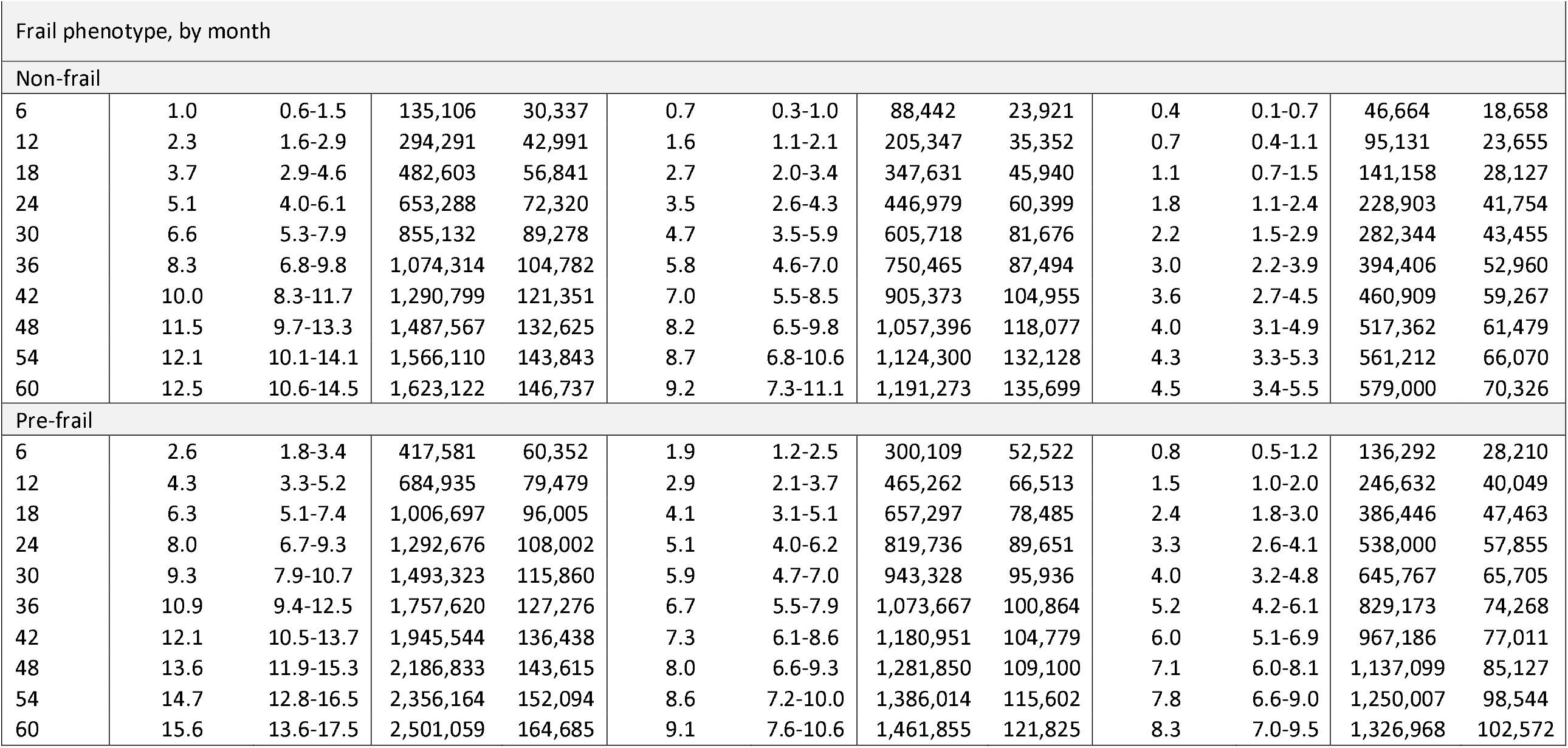
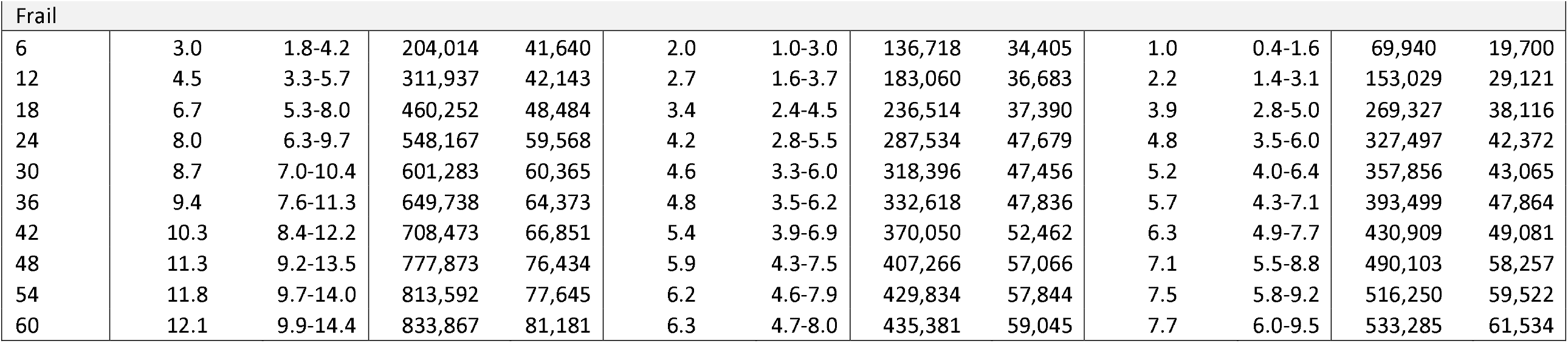
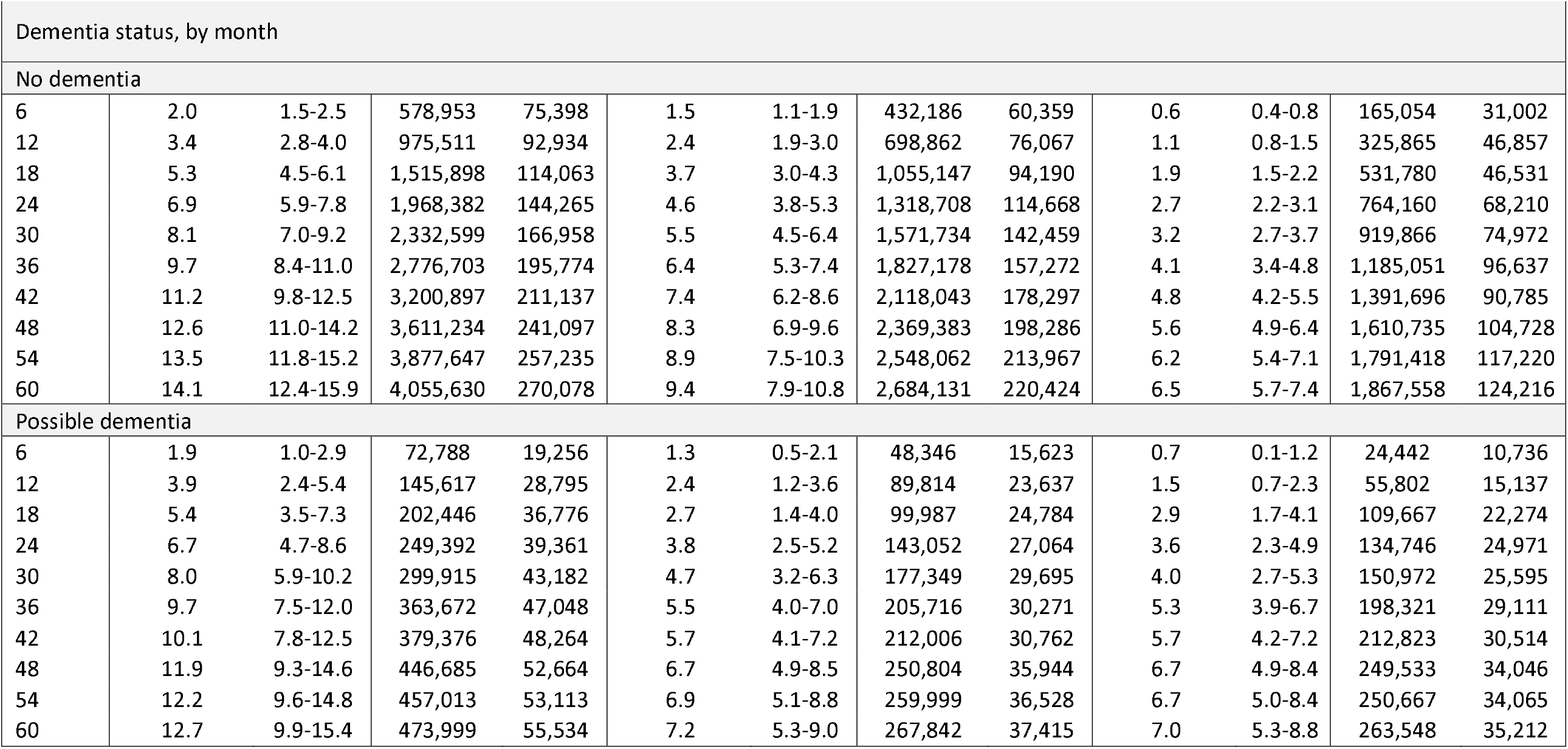
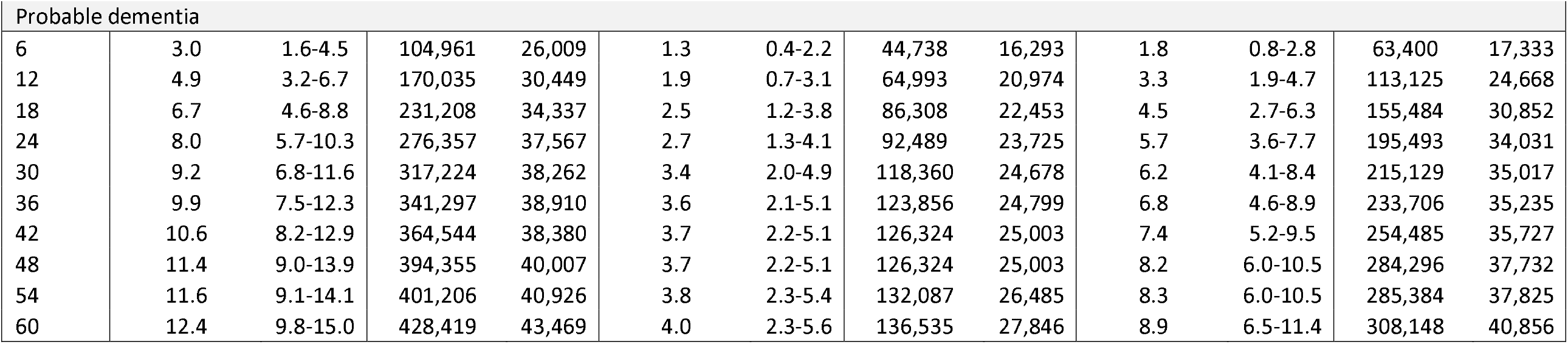
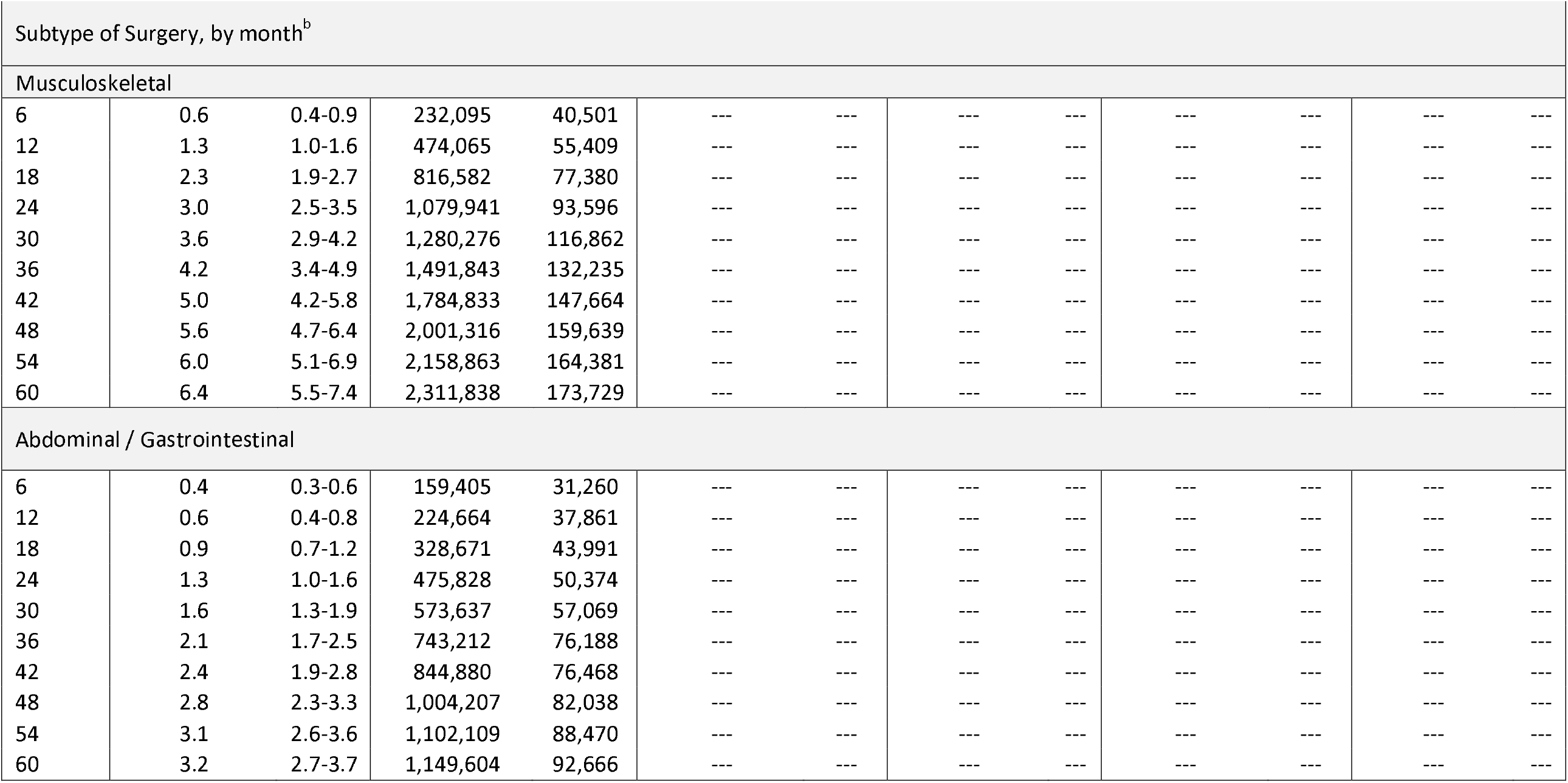
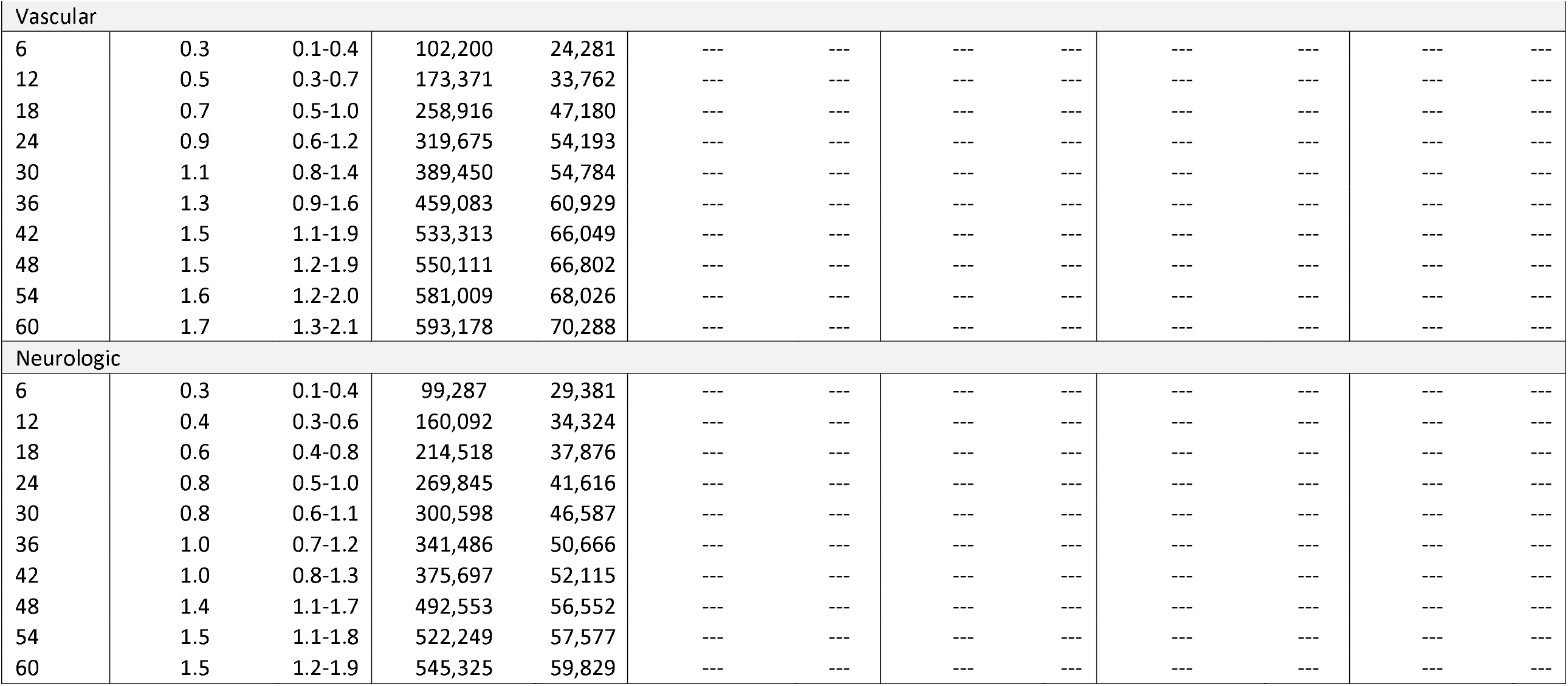
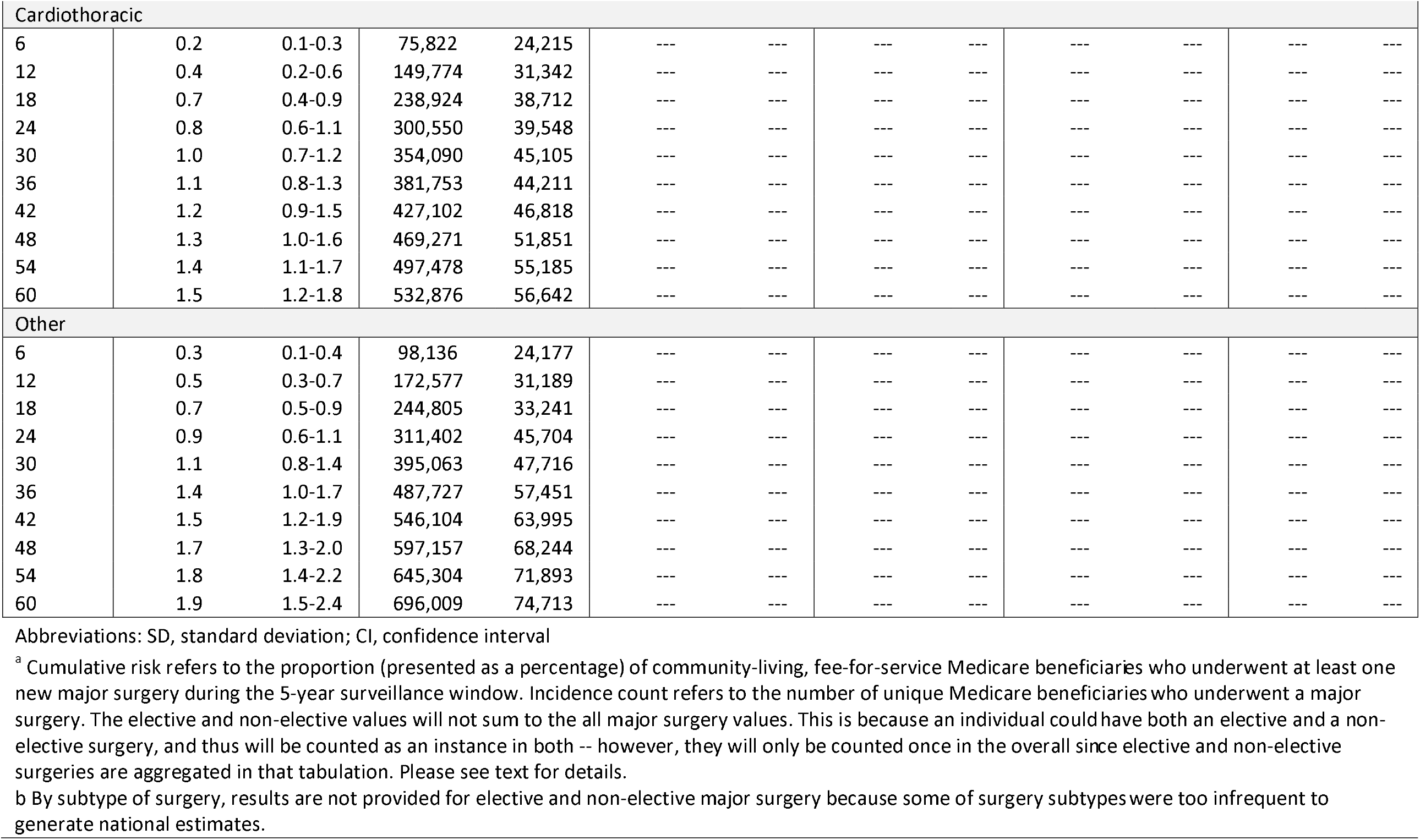
National Estimates of the 5-Year Cumulative Risk of Major Surgery in Community Living Older Adults, by Timing of Operation^a^.

| Variable | All Major Surgery |  |  |  | Elective Major Surgery |  |  |  | Non-Elective Major Surgery |  |  |  |
| --- | --- | --- | --- | --- | --- | --- | --- | --- | --- | --- | --- | --- |
|  | Cumulative Risk | 95% CI | Incidence Count | SD | Cumulative Risk | 95% CI | Incidence Count | SD | Cumulative Risk | 95% CI | Incidence Count | SD |
| Overall risk, by month |  |  |  |  |  |  |  |  |  |  |  |  |
| 6 | 2.1 | 1.6-2.6 | 756,702 | 85,433 | 1.5 | 1.1-1.8 | 525,269 | 68,928 | 0.7 | 0.5-0.9 | 252,896 | 37,071 |
| 12 | 3.6 | 3.0-4.2 | 1,291,163 | 114,199 | 2.4 | 1.9-2.9 | 853,669 | 91,724 | 1.4 | 1.0-1.7 | 494,792 | 59,742 |
| 18 | 5.4 | 4.6-6.2 | 1,949,552 | 140,540 | 3.5 | 2.9-4.1 | 1,241,442 | 110,673 | 2.2 | 1.8-2.6 | 796,931 | 65,089 |
| 24 | 6.9 | 6.0-7.9 | 2,494,131 | 172,596 | 4.3 | 3.6-5.0 | 1,554,249 | 132,910 | 3.0 | 2.6-3.5 | 1,094,400 | 85,616 |
| 30 | 8.2 | 7.1-9.3 | 2,949,738 | 196,806 | 5.2 | 4.3-6.1 | 1,867,443 | 164,927 | 3.6 | 3.1-4.1 | 1,285,967 | 90,114 |
| 36 | 9.7 | 8.5-10.9 | 3,481,672 | 225,125 | 6.0 | 5.1-7.0 | 2,156,750 | 177,082 | 4.5 | 3.9-5.2 | 1,617,077 | 109,207 |
| 42 | 11.0 | 9.7-12.3 | 3,944,816 | 237,948 | 6.8 | 5.8-7.9 | 2,456,374 | 197,250 | 5.2 | 4.6-5.8 | 1,859,004 | 101,784 |
| 48 | 12.4 | 10.9-13.9 | 4,452,274 | 269,736 | 7.7 | 6.5-8.8 | 2,746,511 | 215,755 | 6.0 | 5.3-6.7 | 2,144,564 | 118,082 |
| 54 | 13.2 | 11.6-14.8 | 4,735,867 | 289,504 | 8.2 | 6.9-9.5 | 2,940,147 | 234,757 | 6.5 | 5.7-7.2 | 2,327,469 | 130,812 |
| 60 | 13.8 | 12.1-15.5 | 4,958,048 | 305,858 | 8.6 | 7.3-9.9 | 3,088,509 | 243,445 | 6.8 | 6.0-7.6 | 2,439,253 | 140,781 |
|  | Cumulative Risk | 95% CI | Incidence Count | SD | Cumulative Risk | 95% CI | Incidence Count | SD | Cumulative Risk | 95% CI | Incidence Count | SD |
| Age groups (in years), by month |  |  |  |  |  |  |  |  |  |  |  |  |
| 65-69 |  |  |  |  |  |  |  |  |  |  |  |  |
| 6 | 1.8 | 0.9-2.6 | 192,836 | 44,616 | 1.6 | 0.9-2.4 | 179,064 | 42,248 | 0.1 | 0.0-0.3 | 16,417 | 8,741 |
| 12 | 3.0 | 1.9-4.1 | 330,969 | 60,525 | 2.5 | 1.5-3.5 | 274,941 | 54,380 | 0.5 | 0.2-0.9 | 58,673 | 20,729 |
| 18 | 4.4 | 3.2-5.6 | 479,446 | 65,179 | 3.5 | 2.5-4.6 | 387,111 | 56,827 | 0.9 | 0.5-1.4 | 103,766 | 26,204 |
| 24 | 6.2 | 4.8-7.5 | 675,143 | 77,784 | 4.8 | 3.6-6.0 | 524,755 | 67,659 | 1.8 | 1.1-2.4 | 192,264 | 37,264 |
| 30 | 7.0 | 5.5-8.5 | 763,114 | 85,661 | 5.4 | 4.1-6.8 | 595,839 | 78,044 | 1.9 | 1.2-2.6 | 209,150 | 40,217 |
| 36 | 8.1 | 6.5-9.8 | 892,021 | 95,048 | 6.2 | 4.8-7.6 | 678,307 | 81,148 | 2.5 | 1.7-3.4 | 279,760 | 49,884 |
| 42 | 9.8 | 8.0-11.5 | 1,070,676 | 99,524 | 7.6 | 5.9-9.2 | 828,490 | 91,919 | 2.8 | 2.0-3.7 | 310,875 | 46,381 |
| 48 | 11.5 | 9.6-13.3 | 1,257,078 | 106,199 | 8.6 | 6.9-10.2 | 938,796 | 97,014 | 3.6 | 2.7-4.5 | 398,390 | 49,292 |
| 54 | 12.1 | 10.2-14.0 | 1,327,341 | 113,220 | 9.0 | 7.2-10.8 | 984,561 | 104,405 | 4.0 | 3.0-5.0 | 441,070 | 52,967 |
| 60 | 12.6 | 10.6-14.6 | 1,386,227 | 117,687 | 9.6 | 7.7-11.5 | 1,052,622 | 111,387 | 4.1 | 3.1-5.1 | 450,768 | 55,438 |
| 70-74 |  |  |  |  |  |  |  |  |  |  |  |  |
| 6 | 1.9 | 1.1-2.8 | 158,718 | 34,273 | 1.3 | 0.6-2.0 | 107,597 | 27,517 | 0.7 | 0.3-1.2 | 60,576 | 19,660 |
| 12 | 2.9 | 1.9-3.9 | 237,521 | 39,319 | 2.0 | 1.2-2.8 | 165,826 | 32,989 | 1.1 | 0.5-1.6 | 88,267 | 22,207 |
| 18 | 4.4 | 3.2-5.7 | 363,318 | 52,401 | 2.9 | 1.9-3.8 | 234,499 | 39,738 | 1.8 | 1.0-2.6 | 145,390 | 31,642 |
| 24 | 5.6 | 4.2-7.0 | 460,070 | 59,537 | 3.9 | 2.7-5.0 | 315,976 | 48,104 | 2.1 | 1.3-3.0 | 174,549 | 35,334 |
| 30 | 7.2 | 5.5-8.9 | 588,030 | 73,663 | 4.9 | 3.5-6.3 | 403,446 | 60,002 | 2.7 | 1.8-3.6 | 221,418 | 38,566 |
| 36 | 9.3 | 7.4-11.3 | 765,453 | 84,986 | 5.9 | 4.3-7.4 | 481,352 | 66,278 | 4.2 | 3.0-5.5 | 345,561 | 51,129 |
| 42 | 10.8 | 8.8-12.9 | 886,953 | 90,875 | 6.8 | 5.2-8.5 | 557,401 | 71,838 | 5.3 | 4.1-6.6 | 438,073 | 50,466 |
| 48 | 12.2 | 9.9-14.6 | 1,002,342 | 105,386 | 7.8 | 5.9-9.6 | 634,783 | 81,683 | 6.2 | 4.8-7.5 | 503,922 | 56,874 |
| 54 | 12.9 | 10.4-15.4 | 1,055,302 | 109,266 | 8.6 | 6.6-10.6 | 706,369 | 85,311 | 6.5 | 5.1-7.9 | 535,500 | 59,951 |
| 60 | 13.3 | 10.7-15.9 | 1,090,674 | 114,122 | 8.9 | 6.9-11.0 | 729,625 | 88,379 | 6.8 | 5.4-8.3 | 558,574 | 61,436 |
|  | Cumulative Risk | 95% CI | Incidence Count | SD | Cumulative Risk | 95% CI | Incidence Count | SD | Cumulative Risk | 95% CI | Incidence Count | SD |
| 75-79 |  |  |  |  |  |  |  |  |  |  |  |  |
| 6 | 3.2 | 2.1-4.3 | 204,301 | 36,196 | 2.5 | 1.5-3.4 | 158,093 | 30,028 | 0.8 | 0.3-1.4 | 52,395 | 17,229 |
| 12 | 5.3 | 3.8-6.8 | 337,136 | 46,241 | 4.2 | 3.0-5.3 | 266,004 | 35,199 | 1.5 | 0.7-2.3 | 94,717 | 24,880 |
| 18 | 7.4 | 5.6-9.2 | 471,198 | 56,770 | 5.7 | 4.3-7.2 | 365,314 | 45,243 | 2.2 | 1.2-3.1 | 138,940 | 30,156 |
| 24 | 9.0 | 7.0-10.9 | 570,826 | 59,473 | 6.4 | 4.9-7.9 | 409,816 | 45,750 | 3.2 | 2.1-4.3 | 203,440 | 34,470 |
| 30 | 10.5 | 8.4-12.6 | 671,338 | 63,903 | 7.5 | 5.8-9.2 | 480,865 | 52,028 | 4.0 | 2.9-5.1 | 254,209 | 35,147 |
| 36 | 12.4 | 10.0-14.7 | 787,942 | 69,987 | 8.8 | 6.9-10.6 | 558,106 | 57,117 | 5.1 | 3.8-6.3 | 321,895 | 39,659 |
| 42 | 13.1 | 10.8-15.5 | 837,337 | 71,483 | 9.2 | 7.3-11.1 | 586,237 | 58,460 | 5.6 | 4.3-7.0 | 358,393 | 42,150 |
| 48 | 14.4 | 11.9-16.9 | 917,101 | 74,767 | 9.8 | 7.8-11.8 | 625,233 | 60,250 | 6.3 | 4.9-7.8 | 403,928 | 44,957 |
| 54 | 15.7 | 13.0-18.4 | 1,000,479 | 81,457 | 10.6 | 8.5-12.8 | 676,667 | 64,890 | 7.2 | 5.6-8.8 | 459,393 | 49,222 |
| 60 | 16.6 | 13.8-19.4 | 1,055,385 | 84,528 | 11.2 | 9.0-13.4 | 711,617 | 67,243 | 7.5 | 5.9-9.2 | 480,559 | 49,321 |
| 80-84 |  |  |  |  |  |  |  |  |  |  |  |  |
| 6 | 2.2 | 1.2-3.3 | 113,868 | 26,834 | 0.7 | 0.3-1.1 | 36,447 | 11,038 | 1.6 | 0.6-2.5 | 80,598 | 24,189 |
| 12 | 4.4 | 2.9-6.0 | 229,292 | 40,485 | 1.7 | 1.0-2.4 | 87,081 | 18,570 | 3.0 | 1.5-4.4 | 153,184 | 38,377 |
| 18 | 7.4 | 5.5-9.3 | 384,029 | 51,576 | 3.3 | 2.2-4.4 | 169,948 | 28,136 | 4.4 | 2.9-5.9 | 227,715 | 39,152 |
| 24 | 9.0 | 6.8-11.1 | 463,794 | 57,473 | 3.9 | 2.7-5.1 | 201,429 | 32,242 | 5.5 | 3.9-7.0 | 283,006 | 41,372 |
| 30 | 10.5 | 8.1-12.9 | 542,956 | 65,022 | 5.2 | 3.7-6.7 | 268,760 | 39,099 | 6.1 | 4.4-7.8 | 316,305 | 44,030 |
| 36 | 11.8 | 9.5-14.1 | 608,931 | 62,175 | 5.9 | 4.5-7.3 | 305,150 | 37,481 | 6.8 | 5.2-8.5 | 353,862 | 44,739 |
| 42 | 13.1 | 10.7-15.6 | 679,006 | 65,975 | 6.6 | 5.0-8.1 | 339,835 | 40,440 | 7.8 | 6.0-9.5 | 402,721 | 46,611 |
| 48 | 14.8 | 12.2-17.4 | 765,049 | 69,435 | 7.5 | 5.9-9.2 | 388,589 | 42,026 | 8.7 | 6.8-10.7 | 452,216 | 51,881 |
| 54 | 15.8 | 13.3-18.3 | 816,807 | 69,142 | 7.9 | 6.2-9.5 | 407,680 | 42,390 | 9.4 | 7.5-11.3 | 485,922 | 51,930 |
| 60 | 16.6 | 14.0-19.1 | 856,176 | 70,832 | 8.1 | 6.4-9.7 | 416,724 | 42,685 | 10.0 | 8.0-12.0 | 518,403 | 54,499 |
|  | Cumulative Risk | 95% CI | Incidence Count | SD | Cumulative Risk | 95% CI | Incidence Count | SD | Cumulative Risk | 95% CI | Incidence Count | SD |
| 85-89 |  |  |  |  |  |  |  |  |  |  |  |  |
| 6 | 1.6 | 0.8-2.4 | 53,030 | 12,552 | 0.9 | 0.4-1.5 | 30,537 | 9,159 | 0.7 | 0.2-1.2 | 22,494 | 8,236 |
| 12 | 3.1 | 2.0-4.2 | 102,270 | 16,672 | 1.2 | 0.6-1.8 | 39,975 | 10,401 | 2.0 | 1.1-2.8 | 65,819 | 13,586 |
| 18 | 5.2 | 3.5-7.0 | 173,149 | 28,471 | 1.7 | 1.0-2.4 | 55,952 | 11,375 | 3.7 | 2.1-5.2 | 120,722 | 25,162 |
| 24 | 6.7 | 4.7-8.6 | 221,208 | 32,115 | 2.1 | 1.3-3.0 | 70,439 | 13,212 | 4.8 | 3.2-6.4 | 159,279 | 27,128 |
| 30 | 8.0 | 5.7-10.2 | 263,150 | 36,315 | 2.6 | 1.7-3.6 | 86,697 | 15,745 | 5.6 | 3.8-7.4 | 184,963 | 29,164 |
| 36 | 8.6 | 6.3-11.0 | 285,670 | 37,731 | 3.0 | 1.9-4.1 | 98,654 | 17,754 | 6.0 | 4.2-7.8 | 198,921 | 29,985 |
| 42 | 9.4 | 7.1-11.8 | 312,315 | 37,642 | 3.2 | 2.0-4.3 | 104,482 | 18,655 | 6.6 | 4.8-8.5 | 219,738 | 30,109 |
| 48 | 10.4 | 7.9-12.9 | 344,409 | 40,767 | 3.5 | 2.2-4.8 | 115,909 | 20,536 | 7.6 | 5.5-9.7 | 252,410 | 34,058 |
| 54 | 11.1 | 8.6-13.6 | 367,112 | 41,097 | 3.6 | 2.3-4.9 | 119,138 | 21,309 | 8.2 | 6.1-10.3 | 271,885 | 34,760 |
| 60 | 12.1 | 9.5-14.6 | 398,465 | 41,221 | 3.9 | 2.6-5.3 | 130,103 | 22,381 | 8.8 | 6.7-11.0 | 292,272 | 35,566 |
| 90 and older |  |  |  |  |  |  |  |  |  |  |  |  |
| 6 | 1.8 | 0.6-3.0 | 33,948 | 11,678 | 0.7 | 0.0-1.7 | 13,532 | 8,966 | 1.1 | 0.3-1.9 | 20,417 | 7,481 |
| 12 | 2.9 | 1.4-4.3 | 53,975 | 13,782 | 1.1 | 0.0-2.1 | 19,842 | 9,740 | 1.8 | 0.8-2.8 | 34,133 | 9,522 |
| 18 | 4.2 | 2.6-5.8 | 78,412 | 15,854 | 1.5 | 0.4-2.7 | 28,619 | 11,056 | 3.2 | 1.8-4.7 | 60,398 | 14,434 |
| 24 | 5.5 | 3.9-7.1 | 103,090 | 16,119 | 1.7 | 0.5-2.9 | 31,834 | 11,322 | 4.4 | 3.0-5.8 | 81,862 | 14,511 |
| 30 | 6.4 | 4.6-8.3 | 121,151 | 19,365 | 1.7 | 0.5-2.9 | 31,834 | 11,322 | 5.3 | 3.6-7.1 | 99,922 | 18,391 |
| 36 | 7.5 | 5.6-9.5 | 141,655 | 21,005 | 1.9 | 0.7-3.1 | 35,181 | 11,400 | 6.2 | 4.4-8.1 | 117,079 | 20,170 |
| 42 | 8.4 | 6.4-10.5 | 158,528 | 22,250 | 2.1 | 0.9-3.4 | 39,930 | 11,890 | 6.9 | 4.9-8.8 | 129,204 | 21,196 |
| 48 | 8.9 | 6.9-10.8 | 166,295 | 22,212 | 2.3 | 1.0-3.6 | 43,201 | 12,187 | 7.1 | 5.2-9.0 | 133,699 | 21,016 |
| 54 | 9.0 | 7.1-10.9 | 168,825 | 21,846 | 2.4 | 1.2-3.7 | 45,732 | 11,947 | 7.1 | 5.2-9.0 | 133,699 | 21,016 |
| 60 | 9.1 | 7.2-11.0 | 171,122 | 21,559 | 2.5 | 1.3-3.8 | 47,818 | 12,128 | 7.4 | 5.4-9.3 | 138,677 | 21,252 |
|  | Cumulative Risk | 95% CI | Incidence Count | SD | Cumulative Risk | 95% CI | Incidence Count | SD | Cumulative Risk | 95% CI | Incidence Count | SD |
| Gender, by month |  |  |  |  |  |  |  |  |  |  |  |  |
| Male |  |  |  |  |  |  |  |  |  |  |  |  |
| 6 | 2.7 | 2.0-3.4 | 415,668 | 56,754 | 2.0 | 1.3-2.6 | 303,496 | 51,801 | 0.8 | 0.5-1.2 | 130,992 | 26,087 |
| 12 | 4.1 | 3.2-4.9 | 627,587 | 70,056 | 2.9 | 2.1-3.6 | 442,495 | 60,679 | 1.4 | 1.0-1.8 | 220,739 | 32,630 |
| 18 | 5.7 | 4.5-6.8 | 879,994 | 94,059 | 3.9 | 2.9-4.9 | 604,335 | 81,457 | 2.1 | 1.6-2.6 | 323,436 | 39,629 |
| 24 | 7.1 | 5.8-8.4 | 1,101,010 | 111,977 | 4.7 | 3.5-5.8 | 720,377 | 91,210 | 3.0 | 2.3-3.6 | 459,532 | 54,044 |
| 30 | 8.8 | 7.3-10.2 | 1,359,389 | 122,854 | 5.8 | 4.5-7.1 | 901,478 | 105,835 | 3.5 | 2.8-4.2 | 547,213 | 56,896 |
| 36 | 10.3 | 8.7-12.0 | 1,598,427 | 137,403 | 6.7 | 5.4-8.0 | 1,034,320 | 108,236 | 4.5 | 3.6-5.3 | 690,618 | 64,924 |
| 42 | 11.7 | 10.0-13.3 | 1,806,885 | 137,327 | 7.5 | 6.1-8.9 | 1,163,260 | 112,528 | 5.1 | 4.3-5.9 | 790,591 | 62,349 |
| 48 | 13.0 | 11.2-14.9 | 2,017,589 | 152,053 | 8.4 | 6.9-10.0 | 1,308,196 | 123,017 | 5.7 | 4.8-6.5 | 876,825 | 65,493 |
| 54 | 13.9 | 11.9-15.9 | 2,151,599 | 165,806 | 9.1 | 7.5-10.8 | 1,414,763 | 135,792 | 6.3 | 5.3-7.2 | 968,062 | 71,550 |
| 60 | 14.4 | 12.2-16.5 | 2,228,491 | 176,997 | 9.5 | 7.7-11.2 | 1,468,091 | 142,840 | 6.5 | 5.5-7.5 | 1,006,626 | 77,230 |
| Female |  |  |  |  |  |  |  |  |  |  |  |  |
| 6 | 1.7 | 1.1-2.3 | 341,034 | 60,879 | 1.1 | 0.7-1.5 | 221,774 | 41,968 | 0.6 | 0.3-0.9 | 121,904 | 26,367 |
| 12 | 3.3 | 2.4-4.1 | 663,576 | 85,155 | 2.0 | 1.4-2.6 | 411,174 | 62,762 | 1.3 | 0.9-1.8 | 274,053 | 47,712 |
| 18 | 5.2 | 4.4-6.1 | 1,069,558 | 94,708 | 3.1 | 2.4-3.8 | 637,107 | 73,464 | 2.3 | 1.8-2.8 | 473,495 | 50,406 |
| 24 | 6.8 | 5.8-7.9 | 1,393,121 | 111,173 | 4.1 | 3.3-4.9 | 833,872 | 89,615 | 3.1 | 2.5-3.7 | 634,868 | 58,627 |
| 30 | 7.8 | 6.6-9.0 | 1,590,349 | 124,348 | 4.7 | 3.7-5.7 | 965,964 | 105,304 | 3.6 | 3.0-4.3 | 738,754 | 65,041 |
| 36 | 9.2 | 8.0-10.5 | 1,883,245 | 131,012 | 5.5 | 4.5-6.5 | 1,122,430 | 110,477 | 4.5 | 3.8-5.3 | 926,459 | 73,898 |
| 42 | 10.5 | 9.1-11.8 | 2,137,931 | 145,379 | 6.3 | 5.1-7.5 | 1,293,113 | 128,748 | 5.2 | 4.5-6.0 | 1,068,412 | 77,013 |
| 48 | 11.9 | 10.4-13.5 | 2,434,685 | 163,063 | 7.0 | 5.8-8.3 | 1,438,315 | 134,899 | 6.2 | 5.3-7.1 | 1,267,739 | 89,406 |
| 54 | 12.7 | 11.1-14.2 | 2,584,267 | 168,625 | 7.5 | 6.1-8.8 | 1,525,384 | 142,652 | 6.7 | 5.7-7.6 | 1,359,407 | 93,525 |
| 60 | 13.4 | 11.7-15.0 | 2,729,557 | 174,718 | 7.9 | 6.6-9.3 | 1,620,418 | 144,085 | 7.0 | 6.1-8.0 | 1,432,627 | 97,146 |
|  | Cumulative Risk | 95% CI | Incidence Count | SD | Cumulative Risk | 95% CI | Incidence Count | SD | Cumulative Risk | 95% CI | Incidence Count | SD |
| Frail phenotype, by month |  |  |  |  |  |  |  |  |  |  |  |  |
| Non-frail |  |  |  |  |  |  |  |  |  |  |  |  |
| 6 | 1.0 | 0.6-1.5 | 135,106 | 30,337 | 0.7 | 0.3-1.0 | 88,442 | 23,921 | 0.4 | 0.1-0.7 | 46,664 | 18,658 |
| 12 | 2.3 | 1.6-2.9 | 294,291 | 42,991 | 1.6 | 1.1-2.1 | 205,347 | 35,352 | 0.7 | 0.4-1.1 | 95,131 | 23,655 |
| 18 | 3.7 | 2.9-4.6 | 482,603 | 56,841 | 2.7 | 2.0-3.4 | 347,631 | 45,940 | 1.1 | 0.7-1.5 | 141,158 | 28,127 |
| 24 | 5.1 | 4.0-6.1 | 653,288 | 72,320 | 3.5 | 2.6-4.3 | 446,979 | 60,399 | 1.8 | 1.1-2.4 | 228,903 | 41,754 |
| 30 | 6.6 | 5.3-7.9 | 855,132 | 89,278 | 4.7 | 3.5-5.9 | 605,718 | 81,676 | 2.2 | 1.5-2.9 | 282,344 | 43,455 |
| 36 | 8.3 | 6.8-9.8 | 1,074,314 | 104,782 | 5.8 | 4.6-7.0 | 750,465 | 87,494 | 3.0 | 2.2-3.9 | 394,406 | 52,960 |
| 42 | 10.0 | 8.3-11.7 | 1,290,799 | 121,351 | 7.0 | 5.5-8.5 | 905,373 | 104,955 | 3.6 | 2.7-4.5 | 460,909 | 59,267 |
| 48 | 11.5 | 9.7-13.3 | 1,487,567 | 132,625 | 8.2 | 6.5-9.8 | 1,057,396 | 118,077 | 4.0 | 3.1-4.9 | 517,362 | 61,479 |
| 54 | 12.1 | 10.1-14.1 | 1,566,110 | 143,843 | 8.7 | 6.8-10.6 | 1,124,300 | 132,128 | 4.3 | 3.3-5.3 | 561,212 | 66,070 |
| 60 | 12.5 | 10.6-14.5 | 1,623,122 | 146,737 | 9.2 | 7.3-11.1 | 1,191,273 | 135,699 | 4.5 | 3.4-5.5 | 579,000 | 70,326 |
| Pre-frail |  |  |  |  |  |  |  |  |  |  |  |  |
| 6 | 2.6 | 1.8-3.4 | 417,581 | 60,352 | 1.9 | 1.2-2.5 | 300,109 | 52,522 | 0.8 | 0.5-1.2 | 136,292 | 28,210 |
| 12 | 4.3 | 3.3-5.2 | 684,935 | 79,479 | 2.9 | 2.1-3.7 | 465,262 | 66,513 | 1.5 | 1.0-2.0 | 246,632 | 40,049 |
| 18 | 6.3 | 5.1-7.4 | 1,006,697 | 96,005 | 4.1 | 3.1-5.1 | 657,297 | 78,485 | 2.4 | 1.8-3.0 | 386,446 | 47,463 |
| 24 | 8.0 | 6.7-9.3 | 1,292,676 | 108,002 | 5.1 | 4.0-6.2 | 819,736 | 89,651 | 3.3 | 2.6-4.1 | 538,000 | 57,855 |
| 30 | 9.3 | 7.9-10.7 | 1,493,323 | 115,860 | 5.9 | 4.7-7.0 | 943,328 | 95,936 | 4.0 | 3.2-4.8 | 645,767 | 65,705 |
| 36 | 10.9 | 9.4-12.5 | 1,757,620 | 127,276 | 6.7 | 5.5-7.9 | 1,073,667 | 100,864 | 5.2 | 4.2-6.1 | 829,173 | 74,268 |
| 42 | 12.1 | 10.5-13.7 | 1,945,544 | 136,438 | 7.3 | 6.1-8.6 | 1,180,951 | 104,779 | 6.0 | 5.1-6.9 | 967,186 | 77,011 |
| 48 | 13.6 | 11.9-15.3 | 2,186,833 | 143,615 | 8.0 | 6.6-9.3 | 1,281,850 | 109,100 | 7.1 | 6.0-8.1 | 1,137,099 | 85,127 |
| 54 | 14.7 | 12.8-16.5 | 2,356,164 | 152,094 | 8.6 | 7.2-10.0 | 1,386,014 | 115,602 | 7.8 | 6.6-9.0 | 1,250,007 | 98,544 |
| 60 | 15.6 | 13.6-17.5 | 2,501,059 | 164,685 | 9.1 | 7.6-10.6 | 1,461,855 | 121,825 | 8.3 | 7.0-9.5 | 1,326,968 | 102,572 |
|  | Cumulative Risk | 95% CI | Incidence Count | SD | Cumulative Risk | 95% CI | Incidence Count | SD | Cumulative Risk | 95% CI | Incidence Count | SD |
| Frail |  |  |  |  |  |  |  |  |  |  |  |  |
| 6 | 3.0 | 1.8-4.2 | 204,014 | 41,640 | 2.0 | 1.0-3.0 | 136,718 | 34,405 | 1.0 | 0.4-1.6 | 69,940 | 19,700 |
| 12 | 4.5 | 3.3-5.7 | 311,937 | 42,143 | 2.7 | 1.6-3.7 | 183,060 | 36,683 | 2.2 | 1.4-3.1 | 153,029 | 29,121 |
| 18 | 6.7 | 5.3-8.0 | 460,252 | 48,484 | 3.4 | 2.4-4.5 | 236,514 | 37,390 | 3.9 | 2.8-5.0 | 269,327 | 38,116 |
| 24 | 8.0 | 6.3-9.7 | 548,167 | 59,568 | 4.2 | 2.8-5.5 | 287,534 | 47,679 | 4.8 | 3.5-6.0 | 327,497 | 42,372 |
| 30 | 8.7 | 7.0-10.4 | 601,283 | 60,365 | 4.6 | 3.3-6.0 | 318,396 | 47,456 | 5.2 | 4.0-6.4 | 357,856 | 43,065 |
| 36 | 9.4 | 7.6-11.3 | 649,738 | 64,373 | 4.8 | 3.5-6.2 | 332,618 | 47,836 | 5.7 | 4.3-7.1 | 393,499 | 47,864 |
| 42 | 10.3 | 8.4-12.2 | 708,473 | 66,851 | 5.4 | 3.9-6.9 | 370,050 | 52,462 | 6.3 | 4.9-7.7 | 430,909 | 49,081 |
| 48 | 11.3 | 9.2-13.5 | 777,873 | 76,434 | 5.9 | 4.3-7.5 | 407,266 | 57,066 | 7.1 | 5.5-8.8 | 490,103 | 58,257 |
| 54 | 11.8 | 9.7-14.0 | 813,592 | 77,645 | 6.2 | 4.6-7.9 | 429,834 | 57,844 | 7.5 | 5.8-9.2 | 516,250 | 59,522 |
| 60 | 12.1 | 9.9-14.4 | 833,867 | 81,181 | 6.3 | 4.7-8.0 | 435,381 | 59,045 | 7.7 | 6.0-9.5 | 533,285 | 61,534 |
|  | Cumulative Risk | 95% CI | Incidence Count | SD | Cumulative Risk | 95% CI | Incidence Count | SD | Cumulative Risk | 95% CI | Incidence Count | SD |
| Dementia status, by month |  |  |  |  |  |  |  |  |  |  |  |  |
| No dementia |  |  |  |  |  |  |  |  |  |  |  |  |
| 6 | 2.0 | 1.5-2.5 | 578,953 | 75,398 | 1.5 | 1.1-1.9 | 432,186 | 60,359 | 0.6 | 0.4-0.8 | 165,054 | 31,002 |
| 12 | 3.4 | 2.8-4.0 | 975,511 | 92,934 | 2.4 | 1.9-3.0 | 698,862 | 76,067 | 1.1 | 0.8-1.5 | 325,865 | 46,857 |
| 18 | 5.3 | 4.5-6.1 | 1,515,898 | 114,063 | 3.7 | 3.0-4.3 | 1,055,147 | 94,190 | 1.9 | 1.5-2.2 | 531,780 | 46,531 |
| 24 | 6.9 | 5.9-7.8 | 1,968,382 | 144,265 | 4.6 | 3.8-5.3 | 1,318,708 | 114,668 | 2.7 | 2.2-3.1 | 764,160 | 68,210 |
| 30 | 8.1 | 7.0-9.2 | 2,332,599 | 166,958 | 5.5 | 4.5-6.4 | 1,571,734 | 142,459 | 3.2 | 2.7-3.7 | 919,866 | 74,972 |
| 36 | 9.7 | 8.4-11.0 | 2,776,703 | 195,774 | 6.4 | 5.3-7.4 | 1,827,178 | 157,272 | 4.1 | 3.4-4.8 | 1,185,051 | 96,637 |
| 42 | 11.2 | 9.8-12.5 | 3,200,897 | 211,137 | 7.4 | 6.2-8.6 | 2,118,043 | 178,297 | 4.8 | 4.2-5.5 | 1,391,696 | 90,785 |
| 48 | 12.6 | 11.0-14.2 | 3,611,234 | 241,097 | 8.3 | 6.9-9.6 | 2,369,383 | 198,286 | 5.6 | 4.9-6.4 | 1,610,735 | 104,728 |
| 54 | 13.5 | 11.8-15.2 | 3,877,647 | 257,235 | 8.9 | 7.5-10.3 | 2,548,062 | 213,967 | 6.2 | 5.4-7.1 | 1,791,418 | 117,220 |
| 60 | 14.1 | 12.4-15.9 | 4,055,630 | 270,078 | 9.4 | 7.9-10.8 | 2,684,131 | 220,424 | 6.5 | 5.7-7.4 | 1,867,558 | 124,216 |
| Possible dementia |  |  |  |  |  |  |  |  |  |  |  |  |
| 6 | 1.9 | 1.0-2.9 | 72,788 | 19,256 | 1.3 | 0.5-2.1 | 48,346 | 15,623 | 0.7 | 0.1-1.2 | 24,442 | 10,736 |
| 12 | 3.9 | 2.4-5.4 | 145,617 | 28,795 | 2.4 | 1.2-3.6 | 89,814 | 23,637 | 1.5 | 0.7-2.3 | 55,802 | 15,137 |
| 18 | 5.4 | 3.5-7.3 | 202,446 | 36,776 | 2.7 | 1.4-4.0 | 99,987 | 24,784 | 2.9 | 1.7-4.1 | 109,667 | 22,274 |
| 24 | 6.7 | 4.7-8.6 | 249,392 | 39,361 | 3.8 | 2.5-5.2 | 143,052 | 27,064 | 3.6 | 2.3-4.9 | 134,746 | 24,971 |
| 30 | 8.0 | 5.9-10.2 | 299,915 | 43,182 | 4.7 | 3.2-6.3 | 177,349 | 29,695 | 4.0 | 2.7-5.3 | 150,972 | 25,595 |
| 36 | 9.7 | 7.5-12.0 | 363,672 | 47,048 | 5.5 | 4.0-7.0 | 205,716 | 30,271 | 5.3 | 3.9-6.7 | 198,321 | 29,111 |
| 42 | 10.1 | 7.8-12.5 | 379,376 | 48,264 | 5.7 | 4.1-7.2 | 212,006 | 30,762 | 5.7 | 4.2-7.2 | 212,823 | 30,514 |
| 48 | 11.9 | 9.3-14.6 | 446,685 | 52,664 | 6.7 | 4.9-8.5 | 250,804 | 35,944 | 6.7 | 4.9-8.4 | 249,533 | 34,046 |
| 54 | 12.2 | 9.6-14.8 | 457,013 | 53,113 | 6.9 | 5.1-8.8 | 259,999 | 36,528 | 6.7 | 5.0-8.4 | 250,667 | 34,065 |
| 60 | 12.7 | 9.9-15.4 | 473,999 | 55,534 | 7.2 | 5.3-9.0 | 267,842 | 37,415 | 7.0 | 5.3-8.8 | 263,548 | 35,212 |
|  | Cumulative Risk | 95% CI | Incidence Count | SD | Cumulative Risk | 95% CI | Incidence Count | SD | Cumulative Risk | 95% CI | Incidence Count | SD |
| Probable dementia |  |  |  |  |  |  |  |  |  |  |  |  |
| 6 | 3.0 | 1.6-4.5 | 104,961 | 26,009 | 1.3 | 0.4-2.2 | 44,738 | 16,293 | 1.8 | 0.8-2.8 | 63,400 | 17,333 |
| 12 | 4.9 | 3.2-6.7 | 170,035 | 30,449 | 1.9 | 0.7-3.1 | 64,993 | 20,974 | 3.3 | 1.9-4.7 | 113,125 | 24,668 |
| 18 | 6.7 | 4.6-8.8 | 231,208 | 34,337 | 2.5 | 1.2-3.8 | 86,308 | 22,453 | 4.5 | 2.7-6.3 | 155,484 | 30,852 |
| 24 | 8.0 | 5.7-10.3 | 276,357 | 37,567 | 2.7 | 1.3-4.1 | 92,489 | 23,725 | 5.7 | 3.6-7.7 | 195,493 | 34,031 |
| 30 | 9.2 | 6.8-11.6 | 317,224 | 38,262 | 3.4 | 2.0-4.9 | 118,360 | 24,678 | 6.2 | 4.1-8.4 | 215,129 | 35,017 |
| 36 | 9.9 | 7.5-12.3 | 341,297 | 38,910 | 3.6 | 2.1-5.1 | 123,856 | 24,799 | 6.8 | 4.6-8.9 | 233,706 | 35,235 |
| 42 | 10.6 | 8.2-12.9 | 364,544 | 38,380 | 3.7 | 2.2-5.1 | 126,324 | 25,003 | 7.4 | 5.2-9.5 | 254,485 | 35,727 |
| 48 | 11.4 | 9.0-13.9 | 394,355 | 40,007 | 3.7 | 2.2-5.1 | 126,324 | 25,003 | 8.2 | 6.0-10.5 | 284,296 | 37,732 |
| 54 | 11.6 | 9.1-14.1 | 401,206 | 40,926 | 3.8 | 2.3-5.4 | 132,087 | 26,485 | 8.3 | 6.0-10.5 | 285,384 | 37,825 |
| 60 | 12.4 | 9.8-15.0 | 428,419 | 43,469 | 4.0 | 2.3-5.6 | 136,535 | 27,846 | 8.9 | 6.5-11.4 | 308,148 | 40,856 |
|  | Cumulative Risk | 95% CI | Incidence Count | SD | Cumulative Risk | 95% CI | Incidence Count | SD | Cumulative Risk | 95% CI | Incidence Count | SD |
| Subtype of Surgery, by month<sup>b</sup> |  |  |  |  |  |  |  |  |  |  |  |  |
| Musculoskeletal |  |  |  |  |  |  |  |  |  |  |  |  |
| 6 | 0.6 | 0.4-0.9 | 232,095 | 40,501 | --- | --- | --- | --- | --- | --- | --- | --- |
| 12 | 1.3 | 1.0-1.6 | 474,065 | 55,409 | --- | --- | --- | --- | --- | --- | --- | --- |
| 18 | 2.3 | 1.9-2.7 | 816,582 | 77,380 | --- | --- | --- | --- | --- | --- | --- | --- |
| 24 | 3.0 | 2.5-3.5 | 1,079,941 | 93,596 | --- | --- | --- | --- | --- | --- | --- | --- |
| 30 | 3.6 | 2.9-4.2 | 1,280,276 | 116,862 | --- | --- | --- | --- | --- | --- | --- | --- |
| 36 | 4.2 | 3.4-4.9 | 1,491,843 | 132,235 | --- | --- | --- | --- | --- | --- | --- | --- |
| 42 | 5.0 | 4.2-5.8 | 1,784,833 | 147,664 | --- | --- | --- | --- | --- | --- | --- | --- |
| 48 | 5.6 | 4.7-6.4 | 2,001,316 | 159,639 | --- | --- | --- | --- | --- | --- | --- | --- |
| 54 | 6.0 | 5.1-6.9 | 2,158,863 | 164,381 | --- | --- | --- | --- | --- | --- | --- | --- |
| 60 | 6.4 | 5.5-7.4 | 2,311,838 | 173,729 | --- | --- | --- | --- | --- | --- | --- | --- |
| Abdominal / Gastrointestinal |  |  |  |  |  |  |  |  |  |  |  |  |
| 6 | 0.4 | 0.3-0.6 | 159,405 | 31,260 | --- | --- | --- | --- | --- | --- | --- | --- |
| 12 | 0.6 | 0.4-0.8 | 224,664 | 37,861 | --- | --- | --- | --- | --- | --- | --- | --- |
| 18 | 0.9 | 0.7-1.2 | 328,671 | 43,991 | --- | --- | --- | --- | --- | --- | --- | --- |
| 24 | 1.3 | 1.0-1.6 | 475,828 | 50,374 | --- | --- | --- | --- | --- | --- | --- | --- |
| 30 | 1.6 | 1.3-1.9 | 573,637 | 57,069 | --- | --- | --- | --- | --- | --- | --- | --- |
| 36 | 2.1 | 1.7-2.5 | 743,212 | 76,188 | --- | --- | --- | --- | --- | --- | --- | --- |
| 42 | 2.4 | 1.9-2.8 | 844,880 | 76,468 | --- | --- | --- | --- | --- | --- | --- | --- |
| 48 | 2.8 | 2.3-3.3 | 1,004,207 | 82,038 | --- | --- | --- | --- | --- | --- | --- | --- |
| 54 | 3.1 | 2.6-3.6 | 1,102,109 | 88,470 | --- | --- | --- | --- | --- | --- | --- | --- |
| 60 | 3.2 | 2.7-3.7 | 1,149,604 | 92,666 | --- | --- | --- | --- | --- | --- | --- | --- |
|  | Cumulative Risk | 95% CI | Incidence Count | SD | Cumulative Risk | 95% CI | Incidence Count | SD | Cumulative Risk | 95% CI | Incidence Count | SD |
| Vascular |  |  |  |  |  |  |  |  |  |  |  |  |
| 6 | 0.3 | 0.1-0.4 | 102,200 | 24,281 | --- | --- | --- | --- | --- | --- | --- | --- |
| 12 | 0.5 | 0.3-0.7 | 173,371 | 33,762 | --- | --- | --- | --- | --- | --- | --- | --- |
| 18 | 0.7 | 0.5-1.0 | 258,916 | 47,180 | --- | --- | --- | --- | --- | --- | --- | --- |
| 24 | 0.9 | 0.6-1.2 | 319,675 | 54,193 | --- | --- | --- | --- | --- | --- | --- | --- |
| 30 | 1.1 | 0.8-1.4 | 389,450 | 54,784 | --- | --- | --- | --- | --- | --- | --- | --- |
| 36 | 1.3 | 0.9-1.6 | 459,083 | 60,929 | --- | --- | --- | --- | --- | --- | --- | --- |
| 42 | 1.5 | 1.1-1.9 | 533,313 | 66,049 | --- | --- | --- | --- | --- | --- | --- | --- |
| 48 | 1.5 | 1.2-1.9 | 550,111 | 66,802 | --- | --- | --- | --- | --- | --- | --- | --- |
| 54 | 1.6 | 1.2-2.0 | 581,009 | 68,026 | --- | --- | --- | --- | --- | --- | --- | --- |
| 60 | 1.7 | 1.3-2.1 | 593,178 | 70,288 | --- | --- | --- | --- | --- | --- | --- | --- |
| Neurologic |  |  |  |  |  |  |  |  |  |  |  |  |
| 6 | 0.3 | 0.1-0.4 | 99,287 | 29,381 | --- | --- | --- | --- | --- | --- | --- | --- |
| 12 | 0.4 | 0.3-0.6 | 160,092 | 34,324 | --- | --- | --- | --- | --- | --- | --- | --- |
| 18 | 0.6 | 0.4-0.8 | 214,518 | 37,876 | --- | --- | --- | --- | --- | --- | --- | --- |
| 24 | 0.8 | 0.5-1.0 | 269,845 | 41,616 | --- | --- | --- | --- | --- | --- | --- | --- |
| 30 | 0.8 | 0.6-1.1 | 300,598 | 46,587 | --- | --- | --- | --- | --- | --- | --- | --- |
| 36 | 1.0 | 0.7-1.2 | 341,486 | 50,666 | --- | --- | --- | --- | --- | --- | --- | --- |
| 42 | 1.0 | 0.8-1.3 | 375,697 | 52,115 | --- | --- | --- | --- | --- | --- | --- | --- |
| 48 | 1.4 | 1.1-1.7 | 492,553 | 56,552 | --- | --- | --- | --- | --- | --- | --- | --- |
| 54 | 1.5 | 1.1-1.8 | 522,249 | 57,577 | --- | --- | --- | --- | --- | --- | --- | --- |
| 60 | 1.5 | 1.2-1.9 | 545,325 | 59,829 | --- | --- | --- | --- | --- | --- | --- | --- |
|  | Cumulative Risk | 95% CI | Incidence Count | SD | Cumulative Risk | 95% CI | Incidence Count | SD | Cumulative Risk | 95% CI | Incidence Count | SD |
| Cardiothoracic |  |  |  |  |  |  |  |  |  |  |  |  |
| 6 | 0.2 | 0.1-0.3 | 75,822 | 24,215 | --- | --- | --- | --- | --- | --- | --- | --- |
| 12 | 0.4 | 0.2-0.6 | 149,774 | 31,342 | --- | --- | --- | --- | --- | --- | --- | --- |
| 18 | 0.7 | 0.4-0.9 | 238,924 | 38,712 | --- | --- | --- | --- | --- | --- | --- | --- |
| 24 | 0.8 | 0.6-1.1 | 300,550 | 39,548 | --- | --- | --- | --- | --- | --- | --- | --- |
| 30 | 1.0 | 0.7-1.2 | 354,090 | 45,105 | --- | --- | --- | --- | --- | --- | --- | --- |
| 36 | 1.1 | 0.8-1.3 | 381,753 | 44,211 | --- | --- | --- | --- | --- | --- | --- | --- |
| 42 | 1.2 | 0.9-1.5 | 427,102 | 46,818 | --- | --- | --- | --- | --- | --- | --- | --- |
| 48 | 1.3 | 1.0-1.6 | 469,271 | 51,851 | --- | --- | --- | --- | --- | --- | --- | --- |
| 54 | 1.4 | 1.1-1.7 | 497,478 | 55,185 | --- | --- | --- | --- | --- | --- | --- | --- |
| 60 | 1.5 | 1.2-1.8 | 532,876 | 56,642 | --- | --- | --- | --- | --- | --- | --- | --- |
| Other |  |  |  |  |  |  |  |  |  |  |  |  |
| 6 | 0.3 | 0.1-0.4 | 98,136 | 24,177 | --- | --- | --- | --- | --- | --- | --- | --- |
| 12 | 0.5 | 0.3-0.7 | 172,577 | 31,189 | --- | --- | --- | --- | --- | --- | --- | --- |
| 18 | 0.7 | 0.5-0.9 | 244,805 | 33,241 | --- | --- | --- | --- | --- | --- | --- | --- |
| 24 | 0.9 | 0.6-1.1 | 311,402 | 45,704 | --- | --- | --- | --- | --- | --- | --- | --- |
| 30 | 1.1 | 0.8-1.4 | 395,063 | 47,716 | --- | --- | --- | --- | --- | --- | --- | --- |
| 36 | 1.4 | 1.0-1.7 | 487,727 | 57,451 | --- | --- | --- | --- | --- | --- | --- | --- |
| 42 | 1.5 | 1.2-1.9 | 546,104 | 63,995 | --- | --- | --- | --- | --- | --- | --- | --- |
| 48 | 1.7 | 1.3-2.0 | 597,157 | 68,244 | --- | --- | --- | --- | --- | --- | --- | --- |
| 54 | 1.8 | 1.4-2.2 | 645,304 | 71,893 | --- | --- | --- | --- | --- | --- | --- | --- |
| 60 | 1.9 | 1.5-2.4 | 696,009 | 74,713 | --- | --- | --- | --- | --- | --- | --- | --- |

Abbreviations: SD, standard deviation; CI, confidence interval
<sup>a</sup> Cumulative risk refers to the proportion (presented as a percentage) of community-living, fee-for-service Medicare beneficiaries who underwent at least one new major surgery during the 5-year surveillance window. Incidence count refers to the number of unique Medicare beneficiaries who underwent a major surgery. The elective and non-elective values will not sum to the all major surgery values. This is because an individual could have both an elective and a non-elective surgery, and thus will be counted as an instance in both -- however, they will only be counted once in the overall since elective and non-elective surgeries are aggregated in that tabulation. Please see text for details.
<sup>b</sup> By subtype of surgery, results are not provided for elective and non-elective major surgery because some of surgery subtypes were too infrequent to generate national estimates.

